# Post-Mendelian genetic model in COVID-19

**DOI:** 10.1101/2021.01.27.21250593

**Authors:** Nicola Picchiotti, Elisa Benetti, Chiara Fallerini, Sergio Daga, Margherita Baldassarri, Francesca Fava, Kristina Zguro, Floriana Valentino, Gabriella Doddato, Annarita Giliberti, Rossella Tita, Sara Amitrano, Mirella Bruttini, Laura Di Sarno, Nicola Iuso, Diana Alaverdian, Giada Beligni, Susanna Croci, Ilaria Meloni, Anna Maria Pinto, Chiara Gabbi, Stefano Ceri, Antonio Esposito, Pietro Pinoli, Francis P. Crawley, Elisa Frullanti, Francesca Mari, GEN-COVID Multicenter Study, Marco Gori, Alessandra Renieri, Simone Furini

**Affiliations:** University of Siena, DIISM-SAILAB, Siena, Italy; Department of Mathematics, University of Pavia, Pavia, Italy; Med Biotech Hub and Competence Center, Department of Medical Biotechnologies, University of Siena, Italy; Medical Genetics, University of Siena, Italy; Genetica Medica, Azienda Ospedaliero-Universitaria Senese, Italy; Independent Medical Scientist, Milan, Italy; Politecnico di Milano, DEIB, Milano, Italy; Good Clinical Practice Alliance-Europe (GCPA) and Strategic Initiative for Developing Capacity in Ethical Review-Europe (SIDCER), Leuven, Belgium; Université Côte d’Azur, Inria, CNRS, I3S, Maasai

**Keywords:** COVID-19, Host genetics, Integrative polygenic score

## Abstract

Host genetics is an emerging theme in COVID-19 and few common polymorphisms and some rare variants have been identified, either by GWAS or candidate gene approach, respectively. However, an organic model is still missing. Here, we propose a new model that takes into account common and rare germline variants applied in a cohort of 1,300 Italian SARS-CoV-2 positive individuals. Ordered logistic regression of clinical WHO grading on sex and age was used to obtain a binary phenotypic classification. Genetic variability from WES was synthesized in several boolean representations differentiated according to allele frequencies and genotype effect. LASSO logistic regression was used for extracting relevant genes. We defined about 100 common driver polymorphisms corresponding to classical “threshold model”. Extracted genes were demonstrated to be gender specific. Stochastic rare more penetrant events on about additional 100 extracted genes, when occurred in a medium or severe background (common within the family), simulate Mendelian inheritance in 14% of subjects (having only 1 mutation) or oligogenic inheritance (in 10% having 2 mutations, in 11% having 3 mutations, etc).

The combined effect of common and rare results can be described as an integrated polygenic score computed as: (*n*_*severity*_ − *n*_*mildness*_) + *F* (*m*_*severity*_ − *m*_*mildness*_)

where n is the number of common driver genes, m is the number of driver rare variants and F is a factor for appropriately weighing the more powerful rare variants. We called the model “post-Mendelian”. The model well describes the cohort, and patients are clustered in severe or mild by the integrated polygenic scores, the F factor being calibrated around 2, with a prediction capacity of 65% in males and 70% in females. In conclusion, this is the first comprehensive model interpreting host genetics in a holistic post-Mendelian manner. Further validations are needed in order to consolidate and refine the model which however holds true in thousands of SARS-CoV-2 Italian subjects.

## 1. Introduction

Coronavirus disease 2019 (COVID-19) represents an important testing case for developing new models of complex disorders due to the combination of environmental and genetic factors. Unlike other multifactorial disorders, the main environmental factor can easily be identified through PCR-based tests on swab. Assuming a relatively low impact of viral genome variability [1], the remaining variability might likely be associated with age and host genetics, including gender.

Genome-Wide Association Studies (GWASs) have identified a certain number of common polymorphisms in relevant genes. However, these associations do not fully explain the variability of clinical outcomes [2–3]. The candidate gene approach has shown that, as with many other complex disorders, a simple Mendelian inheritance is also found in COVID-19, affecting some rare individuals with a defect in genes related to innate immunity [4–5].

In a previous study, we explored host genetics through Whole Exome Sequencing analysis (WES) in a cohort of 35 hospitalized COVID-19 patients, which led to the definition of a preliminarily combined model of rare and common variants impacting the clinical outcome [6]. Then, within the Italian GEN-COVID Multicenter Study, biospecimens from more than 1,000 SARS-CoV-2 positive individuals have been collected in the GEN-COVID Biobank (GCB), with clinical data stored in the related Patient Registry (GCPR), and genetic data have in the connected Genetic Data Repository (GCGDR) [7]. SNP genotyping of this cohort contributed to the identification of some loci associated with COVID-19 [3], while WES analysis pinpointed additional common non bi-allelic polymorphisms [8] and rare variants [9]. However, an organic model explaining how common variants may combine with rare variants is still missing. In this study, we propose a new model for predicting the severity of COVID-19 using both common and rare variants. The model was defined in two steps. Initially, logistic regression was used to identify a set of genes that are predictive for the severe or the mild phenotype of COVID-19. Association rules extracted using variants of these genes as input features revealed a robust link between the class of the variants (i.e., protecting or predisposing to the severe phenotype) and the actual phenotype, which supports the predictive capability of the selected genes. In the second step of the model, these predictive genes were used to define a score that separates the extreme COVID-19 phenotypes. This score was able to predict the phenotype in a testing dataset with accuracy of X, proving the predictive capacity of the strategy proposed.

## 2. Materials and methods

### 2.1 Patients cohort and Clinical classification

Demographic and clinical characteristics of the data set were reported in [7]. The number of cases used in this study is 1,318. In order to obtain a clinical classification as independent as possible from age and gender, an Ordered Logistic Regression (OLR) model was used. Separately for the male and female cohorts, two OLR models were fitted using the age to predict the ordinal grading (0, 1, 2, 3, 4) dependent variable **(Figure 1)**. Then, each patient had clinical classification equal to: 0 (mild), if the actual patient grading was below the one predicted by the OLR; or 1 (severe), if the grading was above the OLR prediction. The patients with a predicted gradient equal to the actual gradient were excluded from the LASSO analysis, by which we wanted to compare the “extreme ends”.

**Figure 1.**
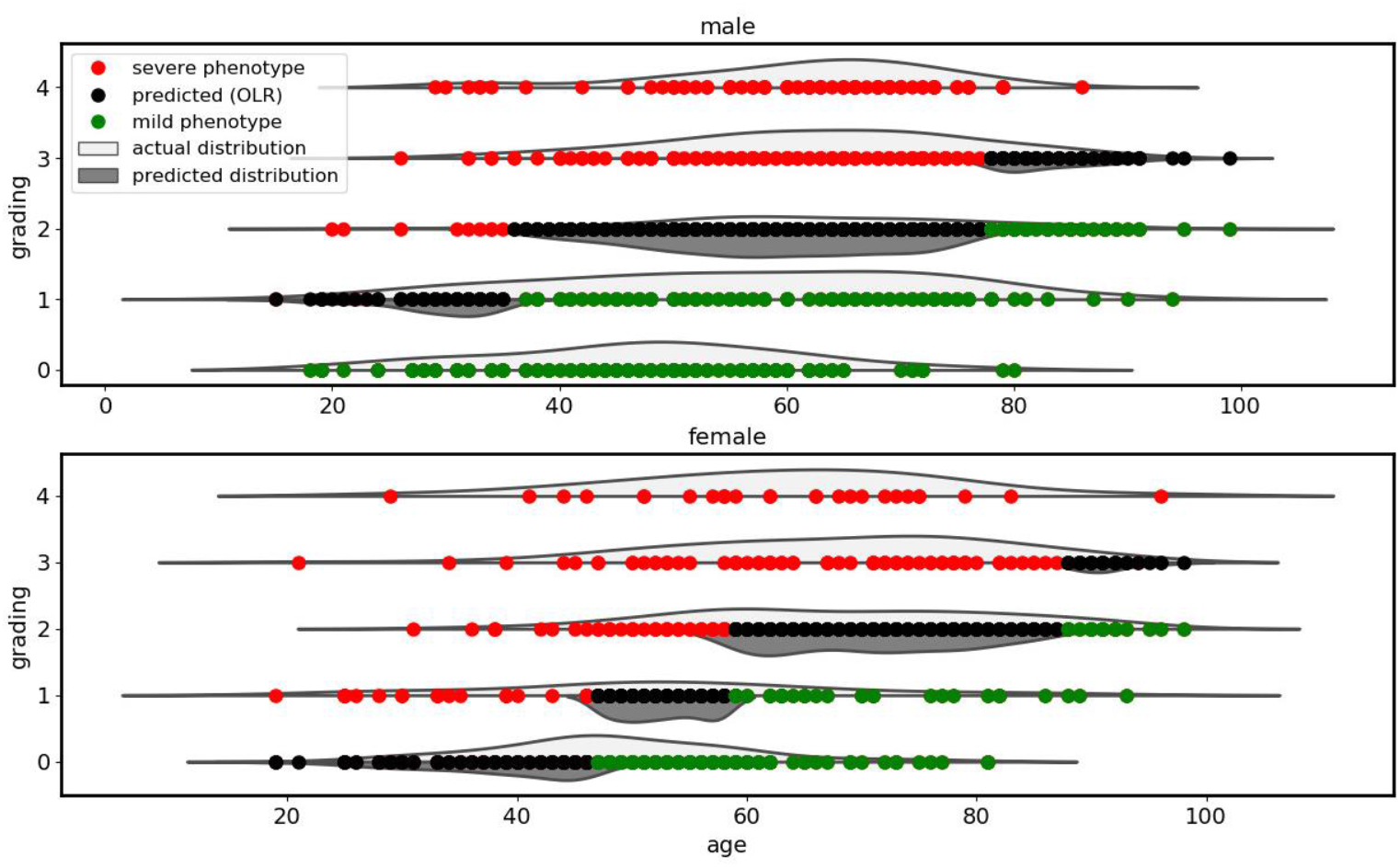
Clinical classification adjusted by age. Two Ordered Logistic Regression (OLR) models, stratified by gender, fitted using the age to predict the ordinal grading (0, 1, 2, 3, 4) dependent variable. On the Y axis, the grading according to patients’ treatment is reported (4=intubated; 3=CPAP/biPAP; 2=oxygen therapy; 1=hospitalized without oxygen support; 0=not hospitalized oligo-asymptomatic patients). On the X axis, age is reported. Red dots represent subjects falling above the expected treatment according to age (hence considered severe), green dots are subjects falling below the expected treatment according to age (hence considered mild) and black dots are subjects matching the expected treatment according to age (hence considered intermediate).

### 2.2 WES analysis

Whole Exome Sequencing with at least 97% coverage at 20x was performed using the Illumina NovaSeq6000 System (Illumina, San Diego, CA, USA). Library preparation was performed using the Illumina Exome Panel (Illumina) according to the manufacturer’s protocol. Library enrichment was tested by qPCR, and the size distribution and concentration were determined using Agilent Bioanalyzer 2100 (Agilent Technologies, Santa Clara, CA, USA). The Novaseq6000 System (Illumina) was used for DNA sequencing through 150 bp paired-end reads. Variants calling was performed according to the GATK4 best practice guidelines, using BWA for mapping and ANNOVAR for annotating. WES data were represented in a binary mode on a gene-by-gene basis.

### 2.3 Boolean representation of bi-allelic polymorphisms

In the boolean representation of bi-allelic polymorphisms, all the unique combinations (mutually exclusive) of common variants (frequency > 1%) with frequency in the cohort above 5% were selected. These unique combinations were used to define a matrix of M by N input features (M and N being respectively the number of combinations and samples), with element j,i equal to 1 if the combination of common variants j is present in sample i.

### 2.4 Boolean representation of rare variants

Three models were proposed for the binary representation of rare variants: autosomal dominant (AD), autosomal recessive (AR), and X-linked (XL). Only coding variants (missense, non-sense, and ins/del), as well as splicing mutations, were considered together with any pathogenic mutations (coding and non-coding) with a frequency below 5% (Phe508del in *CFTR* being 1.1% in the non-Finnish European population). In the AD model, the input feature j,i is 1 if gene j in sample i has at least one variant with a frequency ≤ 1%. In the AR model, the input feature j,i is 1 if gene j in sample i has either a variant with a frequency ≤ 1% in a homozygous state or two variants. In the XL model, only to the male cohort and only genes on chromosomes X are considered. The input feature j,i is 1 if gene j in sample i has a variant with frequency ≤ 1% (hemizygous state).

### 2.5 Sample pre-processing

Among 1,318 samples, after PCA analysis, 63 outliers were removed. Those corresponded to no-white subjects (32 hispanic, 12 black, 18 asian, 1 arabic). In the boolean representation concerning rare variants, genes with higher percentages of mutations than expected as defined by knee analysis were considered artifacts and excluded from the analyses (**Fig. S1)**.

Among the 18,085 annotated genes 12 genes were excluded from the analysis due to error in the annotation such as *ARSD* and *VCX* family gene.

### 2.6 LASSO Logistic Regression

The binary classification problem, i.e., mild/severe cases, was solved by a logistic regression model, one of the most common and successful Machine Learning (ML) 1) common bi-allelic coding haplotypes of autosomal genes (hetero plus homo versus wt) extracting common variants acting in a heterozygous state; 2) common bi-allelic coding haplotypes of autosomal genes (homo versus hetero and wt) extracting common variants acting in a homozygous state; 3) common bi-allelic coding haplotypes of X-linked genes (hemy versus wt) extracting common variants acting in a hemizygous state in males; 4) rare variants of autosomal genes (hetero plus homo versus wt) extracting rare variants acting in heterozygous state; 5) rare variants of autosomal genes (homo versus hetero and wt) extracting rare variants acting in a homozygous state; 6) rare variants of X-linked genes (hemy versus wt) extracting rare variants acting in a hemizygous state in males.algorithms for binary classification tasks with probabilistic interpretation. In order to enforce both the sparsity and the interpretability of the results, the model was trained with the additional LASSO (Least Absolute Shrinkage and Selection Operator) regularization term. By denoting with *β*_*k*_ the coefficients of the logistic regression model and by lambda (λ) the strength of the regularization, the LASSO regularization term of the loss, 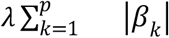, has the effect of shrinking the estimated coefficients to 0, providing a feature selection method for sparse solutions within the classification task. The weights of the logistic regression algorithm can be interpreted as the feature importances of the subset of the most relevant features for the task [10].

The fundamental hyper-parameter of the logistic regression algorithm is the strength of the LASSO term, which was tuned with a grid search method on the average area under the Receiver Operating Characteristic (ROC) curve for the 10-fold cross-validation. 50 equally spaced values in logarithmic scale in the range [10^−3^, 10^2^] were tested. The optimal regularization parameter was selected as the one with the best trade-off between the simplicity of the model and the cross-validation score, i.e., as the highest value providing an average score closer than one standard deviation from the highest score. The rationale of this method is to select the most important genes (and not necessarily the entire set of genes contributing to COVID-19 variability). Data pre-processing was coded in Python, whereas for the logistic regression model, the scikit-learn module with the liblinear coordinate descent optimization algorithm was used.

### 2.7 Association Rules

Association rules are statements in the form “X implies Y”, where, in the current study, X indicates a set of mutated genes and Y indicates the clinical classification of a patient. Association rules can be mined from the data using apriori algorithm. Each association rule is associated with a support, that corresponds to the number of patients for which the antecedent X holds (i.e., patients that have all the genes in the X set mutated) and a confidence, that is the percentage of the patients in the support for which the consequent Y holds. The statistical significance of an association rule can be assessed by comparing the distributions of the values of Y in the set of patients harboring the mutations on the X genes and of the set of all the others, by means of the Fisher’s exact test. In this study, we randomly split the patients in train (80%) and test (20%) sets. We mined the rules on the training set only and used a grid search approach to identify the best pair of support and confidence thresholds. Then, we selected only those rules having a p-value lower than 0.05 on the training set. The remaining rules are further tested on the test set and only those having a Bonferroni corrected p-value lower than 0.05 are kept. For mining association rules we used the arules library (in R).

We mined separately for females and males all significant association rules having support >= 0.08 and confidence >= 0.8, by considering the top genes as extracted by Lasso Logistic Regression. The assessment requires to consider which genes are assembled within rules which associate with either mild or sever course of COVID-19; a confirmation of the model requires genes associated with mild course to be mildness genes, and genes associated with severe course to be severity genes, whereas rules are assembled by our method regardless of a prior assignment to these categories. We also selected enough rules to be associated with high percentages of patients (ranging from 90% of females to 94% of severe cases of males and 99% of mild cases of males).

### 2. 7 The post-Mendelian model

The proposed model extends the standard “threshold model” of common polymorphisms to a more general framework, including the effect of rare variants. The main hypothesis of the model is to describe the combined effect of common and rare results by means of the following “integrated polygenic score” (IPGS):

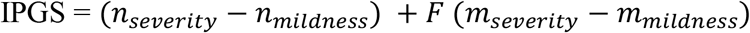

In equation 1, *n*_*severity*_ and *n*_*mildness*_ are the number of common polymorphic driver genes conferring severity or mildness respectively to COVID-19; while *m*_*severity*_ and *m*_*mildness*_ are the number of driver rare variants conferring severity or mildness respectively to COVID-19. The multiplicative factor *F* was included to model a more penetrant effect of rare variants with respect to common variants. In order to identify the optimal value of this weighting factor, for each value of F in the range 0-20, the silhouette coefficient of the clustering (mild vs severe) results was calculated. The optimal value of F was defined as the one that maximizes the silhouette coefficient, i.e., provides the best separation between the two clusters.

## 3. Results

### 3.1 Assessing clinical classification stratified by sex and adjusted by age

Whole Exome Sequencing (WES) data stored in the Genetic Data Repository of the GEN-COVID Multicenter Study (GCGDR) and coming from biospecimens of 1,300 SARS-CoV-2 PCR positive subjects were used for the analysis [7]. Since age and gender are strong determinants of the clinical outcome, we stratified the cohort by gender and then applied ordered logistic regression to re-classify the patients (**Figure 1)** as: i) severe, subjects falling above the expected treatment according to age; ii) intermediate, subjects matching the expected treatment according to age; iii) mild = subjects falling below the expected treatment according to age. This novel classification is expected to be independent of age, and consequently, it should facilitate the identification of the genetic factors responsible for COVID-19 severity.

### 3.2 Discovery of genes and gender dependent effect

We then used severe versus mild cases defined by ordered logistic regression as inputs for a series of logistic regression models for LASSO regularization. The purpose of these logistic models is to identify which features are better predictive for either severity or mildness. Six different types of genetic variability were represented in a boolean manner and tested separately:

For common bi-allelic haplotypes (hetero plus homo versus wt), we identified 19 genes ordered by importance using the cohort with both genders (**Figure S2a**), 12 of them being linked to severity and 7 of them to mildness. Among relevant severity genes are: the haplotype Gly195Glu/Asp213Gly/Glu270Asp of *IBSP*, a sialoprotein part of Coronavirus receptor system, Lys191Thr of *ADAM15*, a negative regulator of TRIF-mediated NF-kB and IFN-b reporter gene activity, Gln5148His/Pro5521Leu of *MUC5AC*, the most frequent mucin in the upper respiratory tract. Among relevant mildness genes are: Ile134Val of *HOXC4*, which enhances antibody response under the regulation of estrogens, and Leu470Val of *DAPK2*, a protein kinase that mediates the body responses to viral infections (**Table S1**).

We then repeated the analysis with gender stratification. We found that the majority of genes, 16 out of 19, are gender-specific, i.e., found in one gender only (specifically 12 in males and 4 in female) (**Figure 4 and Figure S2b and S2c**). For example, the effect of Ile134Val of *HOXC4* was identified in females only, as expected on the base of its biological role, while Gln5148His/Pro5521Leu of *MUC5AC* and Lys191Thr of *ADAM15* was shown to affect males only (**Figure 4**). A similar pattern was found for all the other boolean representations.

The logistic models training with gender stratified cohorts pinpoints an increased number of predictive genes. Indeed, 107 genes were identified in males only and 44 in females only versus 19 in the cohort of both genders. The presence of gender specific effects is the most likely explanation for the increase in the number of predictive genes in gender stratified cohorts (**Figure 2**). Therefore, we decided to proceed with stratification by gender as gender medicine appears to be much more relevant in COVID-19 than in other disorders. In the next two sections, the genes identified in the male and female cohorts are presented.

**Figure 2.**
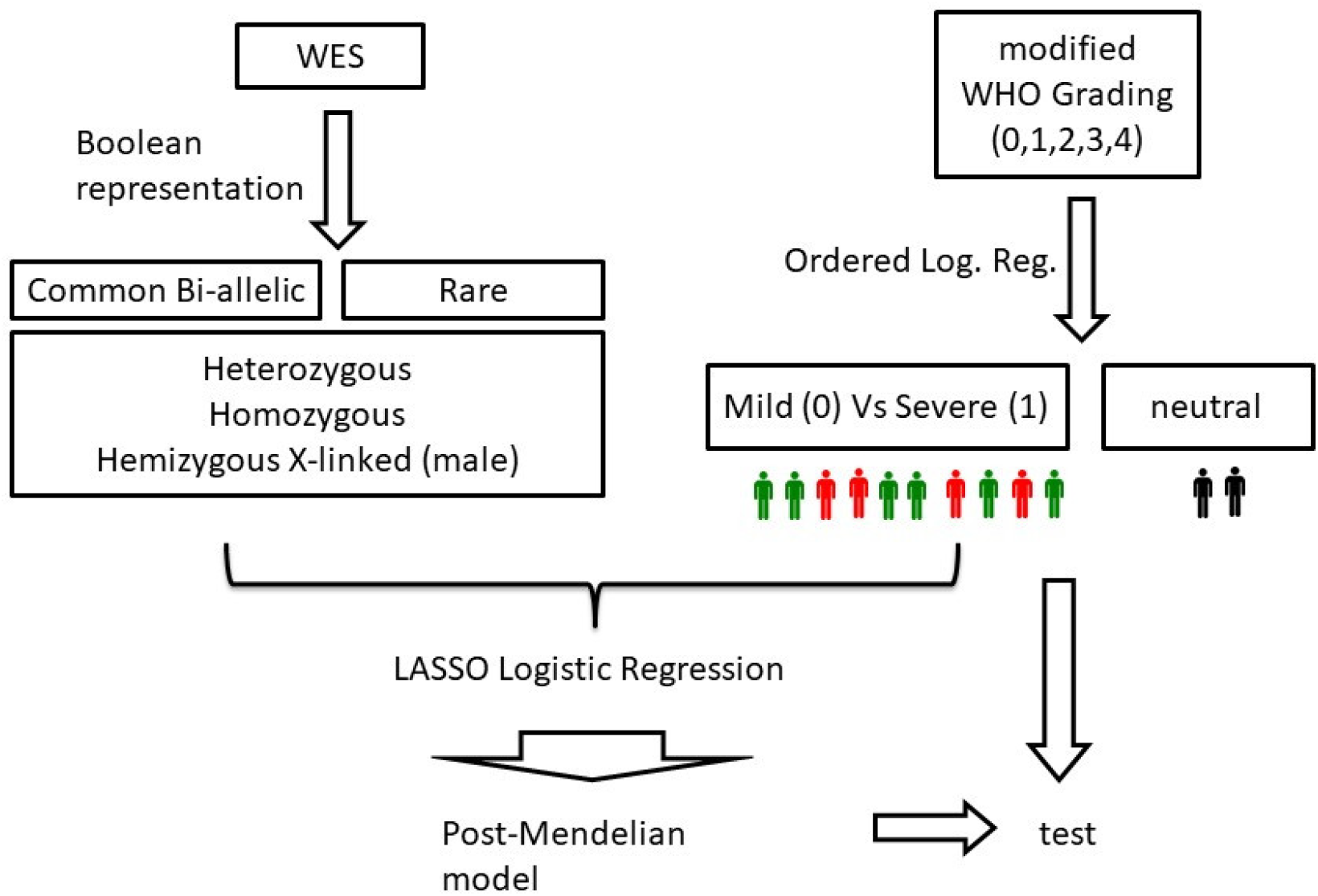
Outline of the method. Separately for the male and female cohorts, two OLR models were fitted using the age to predict the ordinal grading (0, 1, 2, 3, 4) dependent variable. Then, each patient had clinical classification equal to: 0 (green), if the actual patient grading was below the one predicted by the OLR; or 1 (red), if the grading was above the OLR prediction. The patients with a predicted gradient equal to the actual gradient (black) were excluded from the LASSO analysis. LASSO Logistic Regression is performed on WES data represented in a boolean manner, including common and rare variants, separately for males and females. At the end, the post mendelian model is applied. For each subject, an integrated polygenic score (IPGS) is calculated.

**Figure 3.**
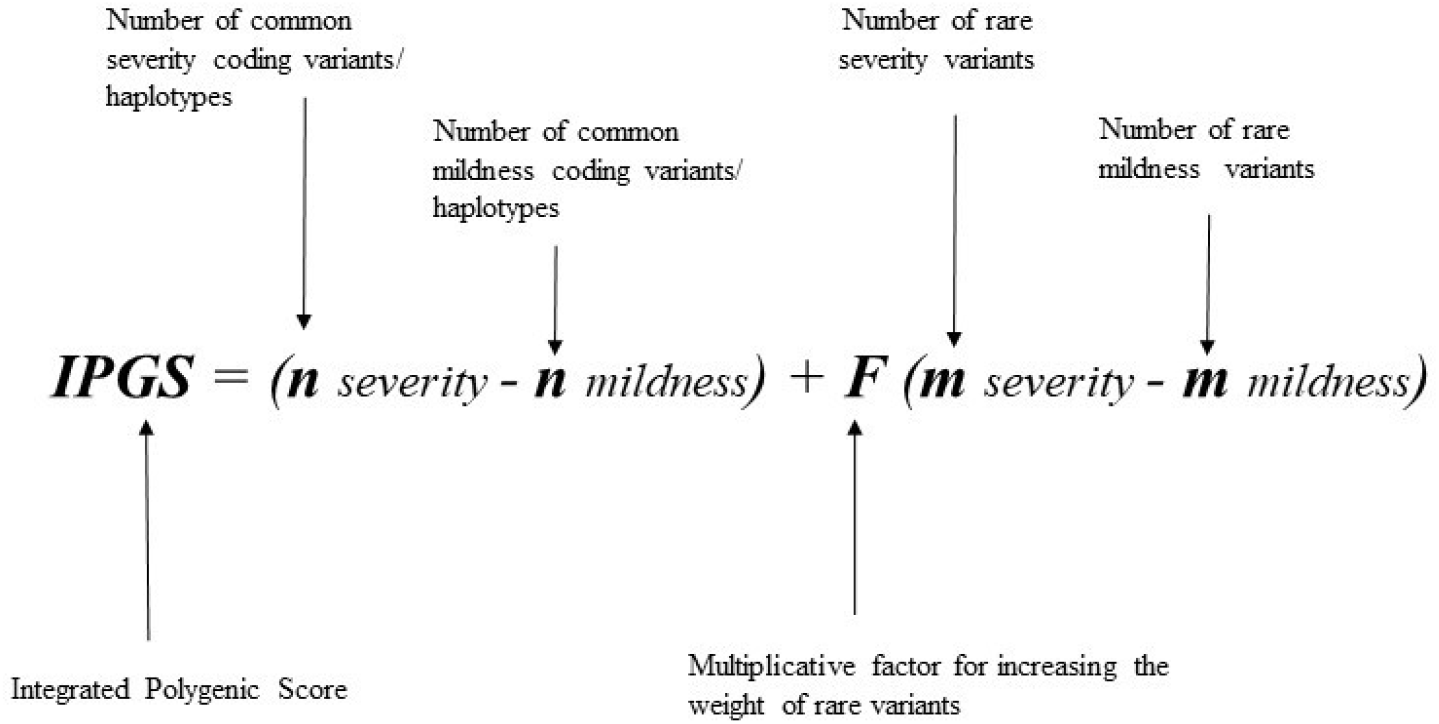
Formula of post-Mendelian model. Formula for the computation of the integrated polygenic score (IPGS). The first term is the difference between the number of common variants/coding haplotypes conferring severity (*n*_*severity*_) and mildness (*n*_*mildness*_) to COVID-19; whereas the second term is the difference between the number of rare variants conferring severity (*m*_*severity*_) and mildness (*m*_*mildness*_) to COVID-19. The multiplicative factor *F* was included to model a more penetrant effect of rare variants with respect to common variants.

### 3.3 Discovery of relevant genes with common and rare variants in males

Using LASSO logistic regression on the boolean representation of common bi-allelic haplotypes and rare variants, we identified 293 relevant genes in males: 154 of them having common variants/haplotypes, while 139 having rare variants.

Among the 154 genes with common variants/haplotypes, in 109, the associated genotype was heterozygous (and homozygous), in 11 homozygous (120 being on autosomes), and in 34 hemizygous, the gene being on the X chromosome. Among the 139 genes with rare variants, in 11, the variant was present in at least one allele (AD-like inheritance), in 69, the variant was present in at least two alleles (AR-like inheritance), and in 59, the variant was hemizygous, the gene being on the X chromosome (XL-like inheritance).

Among relevant extracted **severity** genes with **common** variants are: Val802Ile (hetero) of *C5*, part of the innate immune system, Leu412Phe (hetero) of *TLR3* controlling interferon response, Arg31Pro (hetero) in *TBPL2*, a transcription factor cleaved by Coronavirus protease, Pro285Ala (homo) of *GBP2*, involved in the innate immune system and Glu391Asp (hemi) of *GBC4* gene, a proteoglycan known to be involved in HCV attachment (**Figure S2b, S2e, S2g, and Table S2, S5 and S6**). Among relevant extracted genes with **rare severity** variants are: *MUC5B* (at least 2 variants), a component of mucus secretions, *NEB* (at least 2 variants) involved in inflammatory responses in SARS-CoV-2 infection, and *APOB* (at least 2 variants) apolipoprotein involved in lipid metabolism and HCV hepatocytes entering, *TLR7* (on X Chr) controlling interferon response and *CACNA1F* (on X Chr) calcium channel regulating different processes in T lymphocytes (**Figure S2i, S2n, S2p and Table S8, S11 and S13**).

Among relevant extracted **mildness** genes with **common** variants are: p.Asp611Asn (hetero) of *N4BP2*, required for influenza virus infection and Ile57Val (hetero) of *AURKA*, a cell cycle regulator downregulated during SARS-CoV-2 infection, Ser59Ala (homo) of *RRM2* mildness gene, a cellular factor essential for virus replication, and Arg45His (hemi) of *TXK* mildness gene, involved in immune response (**Figure S2b, S2e, S2g and Table S2, S5, and S6**). Among relevant extracted genes with **rare mildness** variants are: *VPS16* (at least 1 variant), involved in SARS-CoV-2 replication, and *DUOX2* (at least 1 variant), involved in innate immune response *BTNL8* (at least 2 variants), a stimulator of primary immune response, *XPNPEP2* (on X Chr), an *ACE2* network protein involved in SARS-CoV-2 infection and *STK26* (on X Chr), involved immune regulation and inflammatory responses (**Figure S2i, S2n, S2p, and Table S8, S11 and S13**).

We then mined data by association rules in order to identify relevant associations of specific polymorphisms conferring severity: 94% of severe cases of males are covered by the rules reported in **Figure S3a** and **Figure S3b**. Among relevant severity rules in males are *TLR3* Leu412Phe and *IL17RC* Gln267Arg involved in inflammation associated with infection. *MUC5AC* Gln5148His, over expressed in Sars-Cov-2 patients and *TFR*C Gly181Ala, a receptor involved in Sars-cov-2 viral entry (**Figure S3a)**. Among relevant mildness rules in males are *NLRP6* Met163Leu part of inflammasome involved in immune response and *SLC25A5* Leu111Arg a regulator of RNA-viral replication (**Figure S3b**).

### 3.4 Discovery of relevant genes with common and rare variants in females

Using LASSO logistic regression on the boolean representation of common bi-allelic haplotypes and rare variants, we identified 122 relevant genes in females, 78 of them having common variants/haplotypes, while 44 having rare variants. Among the 78 genes with common variants/haplotypes, in 44, the associated genotype was at least heterozygous, in 34 homozygous. Among the 44 genes with rare variants, in 36, the variant was present in at least one allele (AD-like inheritance), in 8, the variant was present in at least two alleles (AR-like inheritance).

Among relevant extracted **severity** genes with **common** variants are: Ile997Val (hetero) of *EP300*, a transcriptional regulator protein involved in chromatin remodeling of proviral genes and Met498Val (hetero) of *HERC5*, involved in antiviral immunity, and Pro33Arg of *TP53* (homo), acting as a host antiviral factor (**Figure S2c and Table S3**).

Among relevant extracted **mildness** genes with **common** variants are: Thr34Ala (hetero) of *HAPLN3*, involved in cell-adhesion and Asn258Ser of *ALCAM* (hetero), a cell adhesion molecule strongly up-regulated in SARS-CoV-2 patients, and Met35Ile (homo) of *APOBEC1*, a deaminase involved in SARS-CoV-2 genome editing (**Figure S2c and Table S3**).

Among relevant extracted **severity** genes with **rare** variants are: *APBA3* (at least 1 variant) involved in antiviral immune responses, *APOL3* (at least one variant), involved in vesicular trafficking and autophagosomes induced by inflammation, *ATG9B* (at least one variant), up-regulated in response to SARS-CoV-2 infection and *SPINT1* (at least 2 variants), associated to SARS-CoV-2 disease severity (**Figure S2l, S2o and Table S9, and S12**).

Among relevant extracted genes with **rare mildness** variants are: *ITGA7* (at least one variant), an integrin involved in post-infection immune response, *MASP1* (at least one variant), a serine protease involved in complement activation, *TNFAIP2* (at least 2 variants), a primary response gene of TNF-alpha. (**Figure S2l, S2o and Table S9, and S12**).

Data mining for association rules reveals that 90% of cases are covered by the rules indicated in **Figure S3c and Figure S3d**. Among severity rules are *TP53* Pro33Arg and *EPCAM* Met 115Thr, an adhesion molecule involved in HBV replication (**Figure S3c**).

Among the mildness rules in females are *APOBEC1* Met80Ile conferring more activity to the RNA editing defence against RNA virus and *CFHR4* Glu125Asp controlling complement (**Figure S3d**).

### 3. 5 The post-Mendelian paradigm for COVID-19 modelization

In the previous sections, the protocol used to identify predictive genes for the severe/mild phenotypes were presented. The plausibility of these genes is supported by biological evidence, and by the fact that association rules for the severe phenotype mostly used genes identified by logistic regression as predisposing to severity, and vice versa.

This agreement among biological knowledge and two independent modelling approaches pushed us to further explore the possibility to combine variants information on these genes into a predictive model of the phenotype. The proposed model extends the standard “threshold model” of common polymorphisms to a more general framework, including both the effect of common and rare variants. In order to discuss the functioning of the model, it will be assumed that the severity of COVID-19 is primarily determined by 200 genes, with approximately half of them presenting common variants, and the remaining rare variants. In case that each variant carries the same relative risk, subjects with approximately the same number of common variants in mildness and severity genes will behave according to their age (black dots in the ordered logistic regression of **Figure 1**).

Instead, subjects with unbalanced variants, that is with a higher number of mildness or severity variants, will more likely belong to the corresponding phenotype, with a probability that increases as the difference between the number of mildness and severity variants increases. Looking for the epidemiological data of the total percentage of those who are severely ill (intubated or CPAP-BiPAP), one should infer the exact number of genes involved in the model and the exact percentage necessary for the threshold effect.

In addition to the cumulative effect of common variants, another group of genes (for simplicity again assumed as composed by 100 genes) may be affected by a rare mutation according to an autosomal dominant, autosomal recessive, or X linked model (with similar probabilities in males). Rare variants have a MAF less than 1%, and like the common variants half of them (about 50) are conferring severity, and half of them (about 50) are conferring mildness when the alternate sequence is present.

Let us suppose a rare mutation occurring in a neutral or severity background of commons. In that case, it will be either a more severe or earlier disease segregating as a Mendelian inheritance (because family members are likely to have identical or similar combinations of common variants). If a rare occurs in a mildness background, the penetrance will depend on the strength of the rare (and likely it is not penetrant).

If a mildness rare occurs in a neutral or mildness background, the individual will be even more mild (for example, an individual with the persistence of antibodies far away from the vaccine or infection). If rare mildness variants occur in a harmful background, its effect will depend on the strength (relevance) of mildness (the type of gene and of mutation) and location of the gene in the pathophysiological process. For example, individuals with rare variants in *FACL4* (which prevents the accumulation of lipid microparticles necessary for the development of the virus) could have milder symptoms even in a negative background (because the action is upstream). On the other hand, for example, there are individuals who are fine after infection but whose swab remains positive for a long time. These could be individuals with common mildness variants but “severity rare mutations” favoring the spread of the virus on the tissues. However the downstream process then appears milder because they respond well by innate and adaptive immunity.

The number of rare variants that each individual can have varies from 0 to more than 6. In individuals without rare variants, the prediction of severity is based on the common variants only. In individuals with rare variants, the correct prediction is made considering common and rares together. In the case of more than one rare in the same sense (e.g., more than one severity rare), the segregation simulates the oligogenic model (digenic, trigenic, etc.).

### 3.6 Fitting the model in the cohort and calibration of rare variants contribution

In the cohort of 1,300 SARS-CoV-2, we tested the above-reported model in which rare variants contribute together with common polymorphisms to COVID-19 liability. For each patient of the cohort, by counting the mutations of the genes extracted by LASSO logistic regression we obtained the integrated polygenic scores reported in the frequency distribution (**Figure 5**) separately for severe (red) and mild (green) phenotypes.

**Figure 4.**
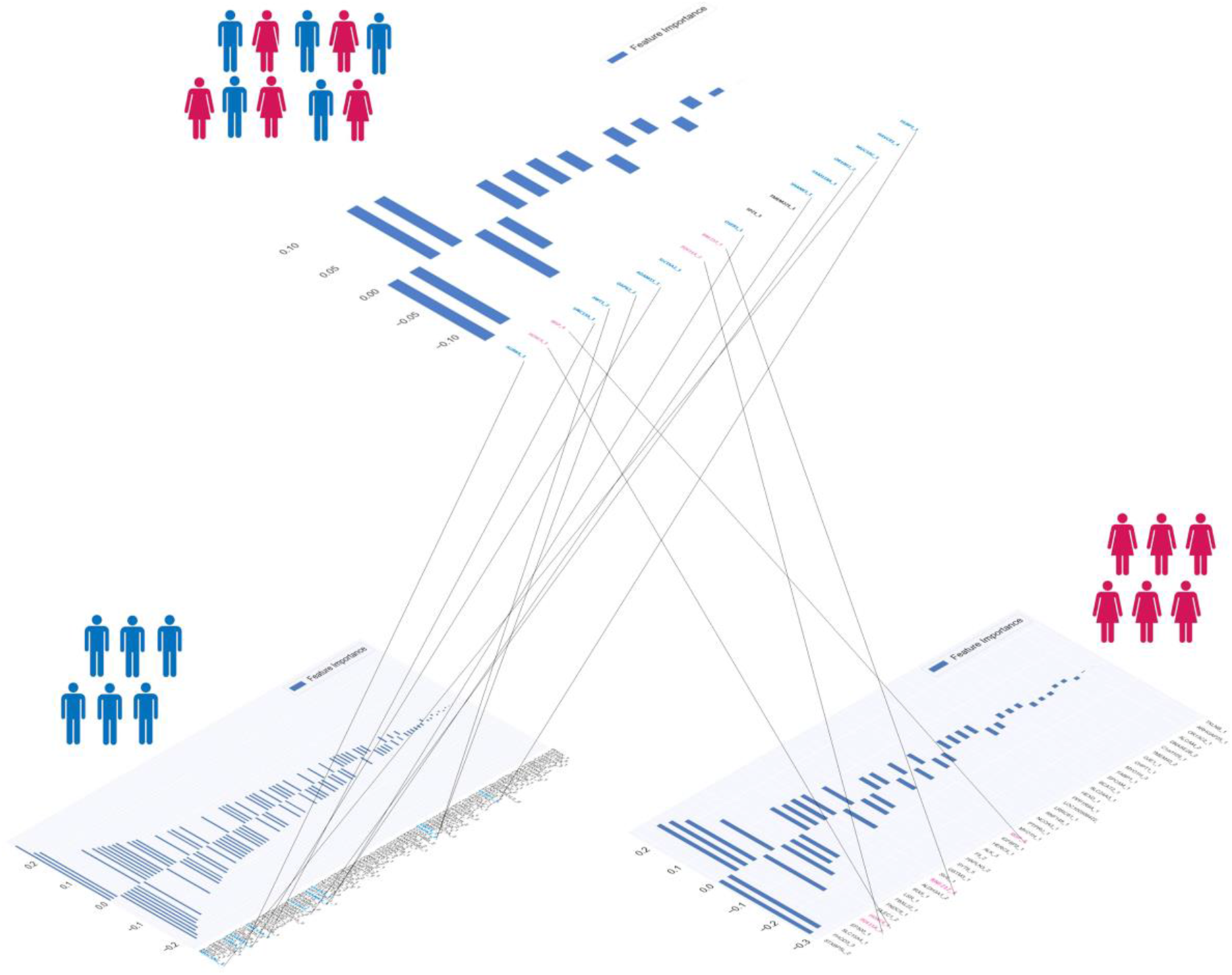
Gender specific effect. **Upper panel:** 19 genes with common bi-allelic haplotypes (hetero plus homo versus wt), ordered by importance in the cohort with both genders. **Down panel**: genes with common bi-allelic haplotypes (hetero plus homo versus wt), ordered by importance and stratifing by gender (left side: 107 genes in males; right side: 44 genes in females). In blue: genes identified only in males. In pink: genes identified only in females.

**Figure 5.**
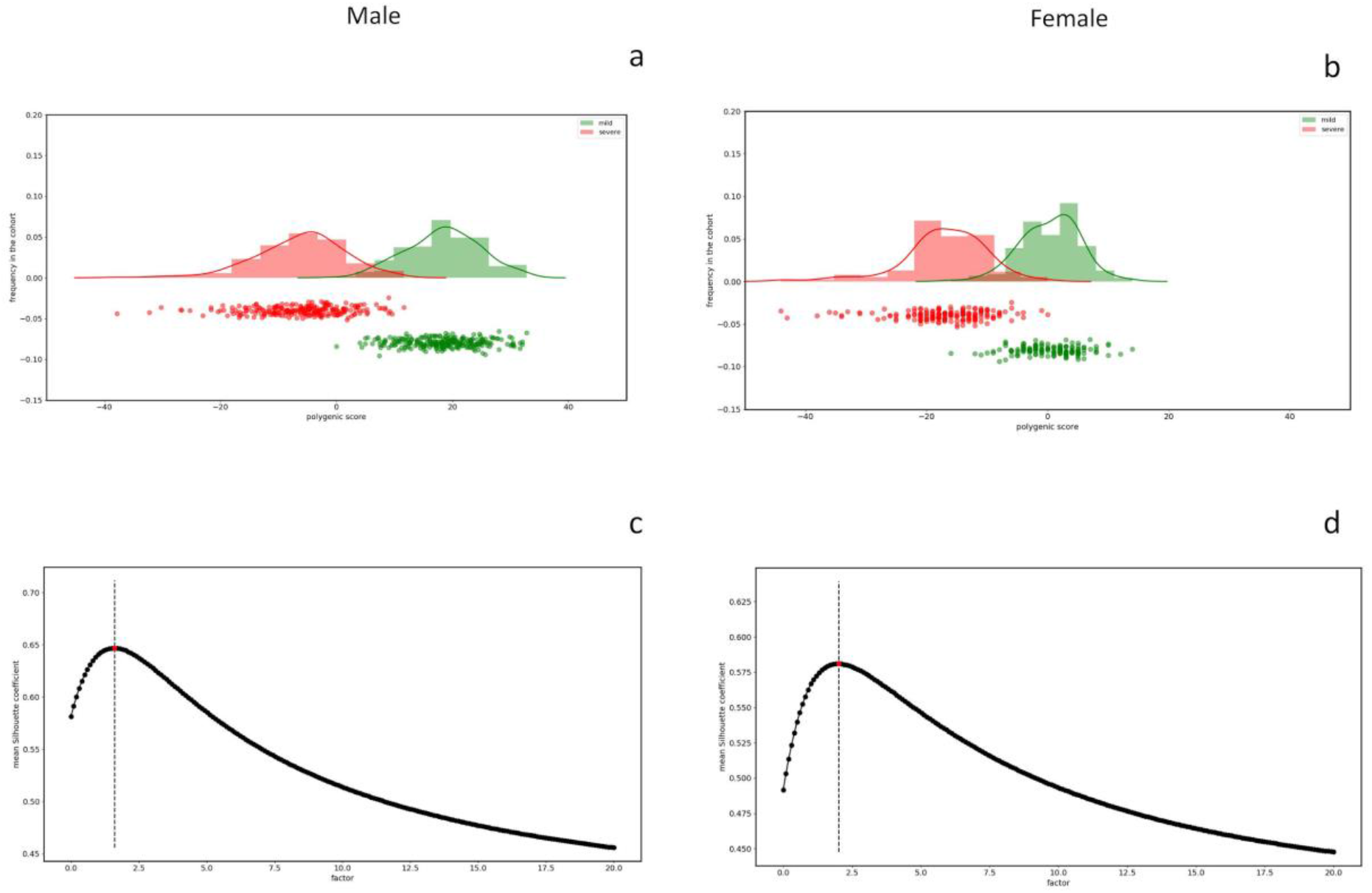
Integrated Polygenic Score, factor calibration and phenotypes. Frequency distributions of the IPGS score for the male cohort (**Panel a**) and female cohort (**Panel b**). The red distribution is related to the severe phenotypes and the green one to the mild phenotypes. **Panel c** and **Panel d** reports the mean silhouette coefficient of the clustering between mild/severe phenotypes as a function of possible F values of the IPGS formula in the range 0-20. The coefficient is a measure of the goodness of the clustering and the maximum value providing the best separation between the two clusters is 1.6 for male (**Panel c**) and 2.1 for female (**Panel d**).

The calibrated F factor, i.e. the coefficient for which the mean silhouette coefficient is maximum, is shown in **Figure 5** for the male (1,6) and female (2,1) cohorts.

In the overall cohort, Mendelian inheritance is simulated when rare variants occurred in a medium or severe background (14% of subjects having only 1 susceptibility mutation) and oligogenic inheritance (10% having 2 susceptibility mutations, 11% having 3 mutations, etc.).

### 3.7 Segregation of the post-Mendelian model in families

In the cohort 15 familial cases were collected and analysed. Segregation analysis using integrated polygenic score perfectly matched the phenotype in all families except one (**Figure 6**). Looking at very rare variants the severely affected member of this family had a very rare pathogenic mutation in *IFNAR1*. The frequency of *IFNAR1* variants is too low to identify them in a cohort of this size by logistic regression.

**Figure 6.**
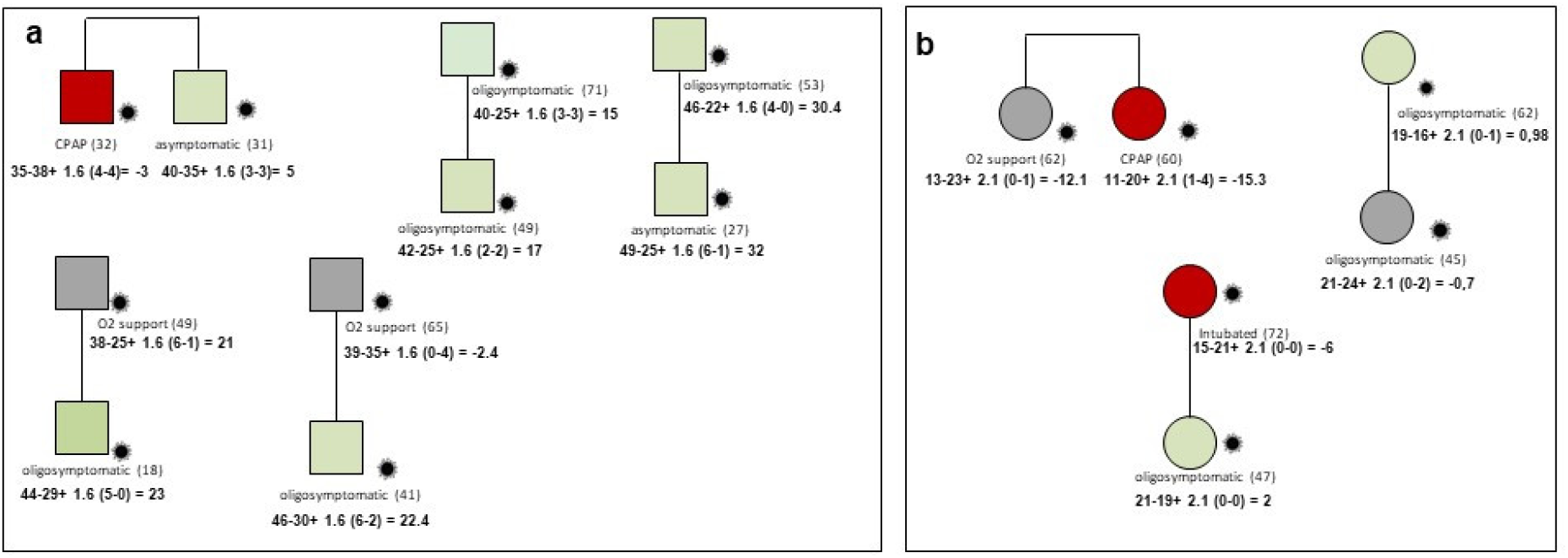
Segregation analysis of post-Mendelian model. Example of segregation analysis of post-Mendelian model using integrated polygenic score in 8 pedigrees. Squares represent severe male and circles females. Red =severely affected (category 1 in fig. 1); green =oligo-asymptomatic subjects (category 0 in fig. 1); grey squares represent intermediate subjects (category black in fig.1). Under symbols is reported the treatment and in parenthesis the age. For each patient is reported the formula for the computation of the integrated polygenic score (IPGS). **Panel a:** Brothers of 32 and 31 years with discordant phenotypes, hospitalized CPAP treated and oligosymptomatic, respectively. In agreement with their phenotype they have IPGS minus 3 and 5, respectively, mainly due to increased severity common polymorphisms in the severely affected brother such as Lys191Thr of *ADAM15* a negative regulator of TRIF-mediated NF-kB and IFN-b reporter gene activity and the polymorphism Asn103Lys in the RNA trafficking gene *RPAIN* which increase viral replication. **Panel b**: Sisters of 62 and 60 years with partially discordant phenotype, hospitalized with oxygen support only and hospitalized CPAP treated, respectively. In agreement with their phenotype they have IPGS minus 12 and minus 15, respectively, mainly due to increased severity of rare variants in the more severely affected sister such as Amyloid Beta Precursor Protein Binding Family A Member 3 *APBA3* which have a role in immune response and low density lipoprotein receptor family member *LRP8* having a role in the suppression of innate response. Treatment with immunosuppressive agents may be an option in this patient.

### 3.8 Testing the model

We repeated the whole analysis by splitting the cohort in a training set (90%) and a testing set (10%) in order to test whether the proposed model is able to generalize the predictions to unseen samples. The total number of genes identified in the training set was 240 in males and 198 in females. Considering only those that were extracted in more than 50% of the 5 repeated cross-validations, we set the number of very relevant genes for both males and females at 135. We therefore tested the predictivity of the model by applying the polygenic score with this set of genes to the testing set. The calibration of F was at 2 for males and at 1 for females. The frequency distributions of the integrated polygenic score are reported in **Figure 7** for the male and the female cohorts. In the upper plots (Panels a and b) the distributions for the training set (90%) are reported whereas the testing set results (10%) are in the lower plots (Panels c and d). The red distribution is related to the severe phenotypes and the green one to the mild phenotypes. Predictivity of the method (ROC-AUC) is 65% and 70%, for males and females respectively.

**Figure 7.**
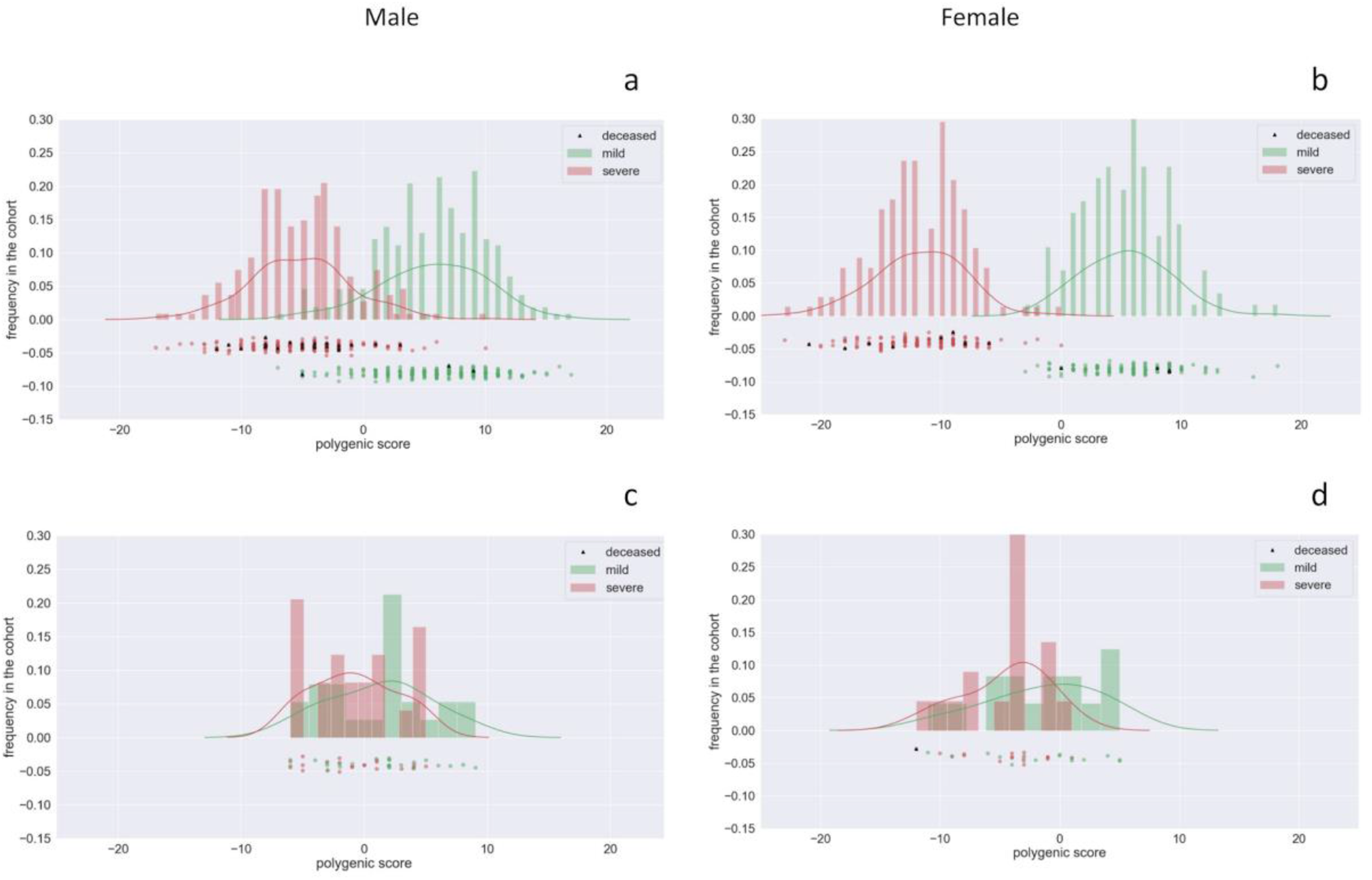
Predictivity of the model. Frequency distributions of the IPGS score for the male cohort (**Panels a** and **c**) and female cohort (**Panel b** and **d**). In the upper plots the distributions of integrated polygenic score and phenotype distribution for the training set (90%) are reported; in the lower plot the distribution for the testing set (10%) is reported. The red distribution is related to the severe phenotypes and the green one to the mild phenotypes. The predictivity is 65% for males and 70% for females.

## 4. Discussion

Using machine learning methodologies, we identified a subset of genes predisposing to severe COVID-19 and a subset of genes preventing the development of such a disease, thus conferring a mildness effect (**Figure 2**). Most importantly, we defined a new genetic model for explaining genetic susceptibility to COVID-19 (**Figure 3**). The proposed method has a number of innovative approaches.

First, the clinical modified WHO outcome scale was combined with the age, that is the main “non genetic” factor influencing clinical outcome, dividing subjects in three categories only: those having a clinical outcome as expected just for age and those having either a worse or better clinical outcome. The last two were selected as extreme ends of phenotype and used for selecting relevant genes.

Second, the method is gene based and processes the genes in a Boolean manner, i. e. having or not having that variant. For both common and rare variants the Boolean classification was repeated three times corresponding of having at least the heterozygous genotype (dominant modes), homozygous genotype (recessive model) or hemizygous genotype (X-linked model) for genes on chromosome X and males only.

Third, we reasoned that not rarely (3413) genes have more than one coding polymorphism. All the coding changes may move the function of the protein either in the same direction (for example lowering or increasing the function) or in the opposite direction therefore cancelling each other. We used a “coding haplotype” approach for classifying polymorphic variability in a Boolean manner: we counted the most common combinations (not all theoretical combinations are present due to linkage disequilibrium) and assigned the score of 1 to each gene if the specific combination is present or zero if not. Again the Boolean classification was repeated three times corresponding to having at least the heterozygous genotype, homozygous genotype or hemizygous genotype for genes on chromosome X and males only.

Fourth, the method is treating separately males and females thus representing the first approach fully translating into practice gender medicine. Indeed, male patients affected by COVID-19 undergo a more severe clinical course.

Fifth, the method we used for gene selection, based on Lasso regularization for a logistic regression classifier, is indirectly confirmed by association rules, produced by a different method, as protection genes spontaneously associate with mild cases and severity genes associate with severe cases.

Finally, the method is able to take into account both common and rare variability. To our knowledge this is the first method able to synthesise in an holistic approach both variabilities, which usually are treated separately: using polygenic score from GWAS approach for the common variability and Mendelian model for rare variability. We therefore suggest to call this method “post-Mendelian model”.

However, the method is still far from perfect and needs further improvement. For example the method is treating each gene as having the same weight within common or within rare variants.

Furthermore, a number of genetic variabilities are missed in the model such as: i) non biallelic common polymorphisms like polyamino acid repeats, known to contribute to COVID-19 [8]; ii) variability due to germline CNV; iii) variability due to somatic mutations, both CNV and SNP [12–13]; iv) very rare variants or private variants, whose frequency is not enough to be detected by LASSO logistic regression.

Further studies are needed in order to improve the model taking into account the above reported variability and increase prediction performances.

## 5. Conclusion

We were able to identify a set of genes in which common polymorphisms confer either severity or mildness against COVID-19 severity. We also identified another set in which rare variants confer either severity or mildness to COVID-19 disease. We have finally defined a new post-Mendelian genetic model, based on an analysis of common and rare variants, that can explain severity in COVID-19.

The model is good enough to extract the relevant genes of a specific patient and understand the main pathogenic pathways useful for assessing a personalized co-adjuvant treatment flanking the current use of cortisone. It is still young to be used as a predictive tool in not yet infected individuals, but it represents the basis for a second release of the model with potential predictive use.

By better understanding the role of host genetics in COVID-19 susceptibility, we are also in a stronger position to identify public health measures that will help to curb the impact of the disease on society as a whole. This should help us to genetically screen already affected patients as well as individuals who may potentially be patients in order to predict those who are more or less susceptible to developing COVID-19 post-infection. It should further help us in, not only reassigning therapeutics or developing new interventions (including vaccines), but also in decision-making regarding therapeutics and vaccine allocations. Beyond what this post-Mendelian method can help us understand regarding the role of host genetics in COVID-19 susceptibility and the potential implications for clinical and public health responses, the model also has strong potential for understanding the role of host genetics in other complex disorders, as alluded to above. As we move into an era of precision, patient-centric medicine, this post-Mendelian method can help us tailor treatments to the specific needs of individual patients.

## Data Availability

The data and samples referenced here are housed in the GEN-COVID Patient Registry and the GEN-COVID Biobank and are available for consultation. You may contact the corresponding author, Prof. Alessandra Renieri.

## FUNDINGS

MIUR project “Dipartimenti di Eccellenza 2018-2020” to Department of Medical Biotechnologies University of Siena, Italy (Italian D.L. n.18 March 17, 2020). Private donors for COVID-19 research. “Bando Ricerca COVID-19 Toscana” project to Azienda Ospedaliero-Universitaria Senese. Charity fund 2020 from Intesa San Paolo dedicated to the project N. B/2020/0119 “Identificazione delle basi genetiche determinanti la variabilità clinica della risposta a COVID-19 nella popolazione italiana”.

## ACKNOWLEDGMENTS

This study is part of the GEN-COVID Multicenter Study, https://sites.google.com/dbm.unisi.it/gen-covid, the Italian multicenter study aimed at identifying the COVID-19 host genetic bases. Specimens were provided by the COVID-19 Biobank of Siena, which is part of the Genetic Biobank of Siena, member of BBMRI-IT, of Telethon Network of Genetic Biobanks (project no. GTB18001), of EuroBioBank, and of D-Connect. We thank the CINECA consortium for providing computational resources and the Network for Italian Genomes (NIG) http://www.nig.cineca.it for its support. We thank private donors for the support provided to A.R. (Department of Medical Biotechnologies, University of Siena) for the COVID-19 host genetics research project (D.L n.18 of March 17, 2020). We also thank the COVID-19 Host Genetics Initiative (https://www.covid19hg.org/).

## AUTHOR CONTRIBUTIONS

Conceptualization, Elisa Frullanti, Francesca Mari and Alessandra Renieri; Data curation, Chiara Fallerini, Marco Gori and Stefano Ceri; Formal analysis, Nicola Picchiotti, Elisa Benetti, Chiara Fallerini, Sergio Daga, Margherita Baldassarri, Francesca Fava, Kristina Zguro, Stefano Ceri, Antonio Esposito, Pietro Pinoli and Simone Furini; Funding acquisition, Alessandra Renieri; Methodology, Floriana Valentino, Gabriella Doddato, Annarita Giliberti, Sara Amitrano and Mirella Bruttini; Project administration, Alessandra Renieri; Supervision, Ilaria Meloni, Alessandra Renieri and Simone Furini; Validation, Alessandra Renieri and Simone Furini; Visualization, Elisa Benetti, Chiara Fallerini, Sergio Daga, Laura Di Sarno, Nicola Iuso, Diana Alaverdian and Susanna Croci; Writing – original draft, Nicola Picchiotti, Elisa Benetti, Chiara Fallerini, Sergio Daga, Margherita Baldassarri, Francesca Fava, Kristina Zguro, Floriana Valentino, Gabriella Doddato, Annarita Giliberti, Rossella Tita, Sara Amitrano, Mirella Bruttini, Ilaria Meloni, Anna Maria Pinto, Chiara Gabbi, Francis P. Crawley, Elisa Frullanti, Francesca Mari, GEN-COVID Multicenter Study, Marco Gori, Stefano Ceri, Antonio Esposito, Pietro Pinoli, Alessandra Renieri and Simone Furini.

## INSTITUTIONAL REVIEW BOARD STATEMENT

The GEN-COVID study was conducted according to the guidelines of the Declaration of Helsinki, and approved by the University Hospital of Siena Ethical Review Board (protocol code 16929, dated March 16, 2020).

## INFORMED CONSENT STATEMENT

The patients were informed of this research and agreed to it through the informed consent process.

## CONFLICTS OF INTEREST

The authors declare no conflict interests.

## ETHICS APPROVAL

The GEN-COVID study was approved by the University Hospital of Siena Ethical Review Board (Protocol n. 16929, dated March 16, 2020).

## SUPPLEMENTARY MATERIALS

**Figure S1. Knee analysis**

In the boolean representation of rare variants, the genes with a number of mutations higher than a threshold are excluded from the LASSO analysis. The threshold has been defined by the knee-analysis of the empirical cumulative density function of the number of mutations for the genes in the overall cohort.

**Figure S2. Common polymorphisms and rare variants selected by LASSO Logistic Regression.**

Comparison of extreme ends of phenotype: subjects of class 1 (red in Fig.1) versus class 0 (green in Fig. 1) as defined by ordered logistic regression.

Common (>1%) bi-allelic polymorphisms (coding haplotypes) considering heterozygous variants plus homozygous variants versus wt genotype and using both males and females (**S2a**) or only males (**S2b**) or only females (**S2c**); Common (>1%) bi-allelic polymorphisms (coding haplotypes) considering homozygous variants versus heterozygous variants plus wt genotype and using both males and females (**S2d**) or only males (**S2e**). For females (**S2f**) the performances are below the random guess. Common (>1%) bi-allelic polymorphisms (coding haplotypes) considering hemizygous variants (only genes on chromosome X) versus wt genotype and using males only (**S2g**);

Rare variants (<1%) considering considering heterozygous variants plus homozygous variants versus wt genotype (**dominant model**) and using both males and females (**S2h**) or only males (**S2i**) or only females (**S2l**); Rare variants (<1%) considering homozygous variants versus heterozygous variants plus wt genotype (**recessive model**) and using both males and females (**S2m**) or only males (**S2n**) or only females (**S2o**); Rare variants (<1%) considering hemizygous variants (only genes on chromosome X) versus wt genotype (**X-linked model**) and using males only (**S2p**). Features are gene-based representations of Genotypic Combinations (GC) of common polymorphisms. The histograms (weights) represented by importance of each feature (genes), inlcuding age and sex, for the classification task (**Upper Panel**). The positive weights reflect a susceptible behaviour of the gene to the target COVID-19 disease, whereas the negative weights a mildness action. **Down Panel**: Cross-validation accuracy for the grid of LASSO regularization parameters; the error bar is given by the standard deviation of the average ROC-AUC within the 10 folds; the red point corresponds to the parameter chosen for the fitting procedure.

**Figure S3. Model assessment by means of association rules**

We ordered the rules, each typically assembling four to five genes, by decreasing support. We then built curves of cumulative counters of distinct patients associated with at least one such rule; we considered as preferred cutting point of each curve a point at the start of a “plateau” such that cumulative counter is relative to more than 90% of patients (actually, 90% of females, 94% of severe cases of males and 99% of mild cases of males). The most remarkable aspect of this construction, obtained by a data mining method which is completely different from the ML-based method used for gene selection, is that genes assembled within mined rules are “coherent”: both for males and females, mildness genes associate with mild cases and severity genes associate with severe cases. Out of several hundred genes included in the rules, only four are “incoherent” (marked in yellow). They are also widely mutated and hence not really significant, as genes *PRSS5, APOBEC1* and *SLC24A3* are mutated respectively in 55%, 75% and 86% of females, and gene *FOXR2* is mutated in 94% of males. **Panel a: Male/Severity**. Selected rules are significant (P<0.05), have support>0.08 and confidence>0.8, ordered by support (also reported as count of matching patients); confidence is illustrated by a graded scale. Top rules are selected based on the cumulative number of distinct patients associated with at least one rule (99%).

Remarkably, all genes of top rules are severity genes. **Panel b: Male/Mildness**. Selected rules are significant (P<0.05), have support>0.08 and confidence>0.8, ordered by support (also reported as count of matching patients); confidence is illustrated by a graded scale. Top rules are selected based on the cumulative number of distinct patients associated with at least one rule (99%). Remarkably, all genes of top rules are mildness genes except gene FOXR2, which however is present in 94% of male patients. **Panel c**: **Female/Severity**.

Selected rules are significant (P<0.05), have support>0.08 and confidence>0.8, ordered by support (also reported as count of matching patients); confidence is illustrated by a graded scale. Top rules are selected based on the cumulative number of distinct patients associated with at least one rule (90%). Remarkably, all genes of top rules are mildness genes except genes APOBEC1 and SLC24A3 (present respectively in 86% and 75% of female patients). **Panel d: Female/Mildness**. Selected rules are significant (P<0.05), have support>0.08 and confidence>0.8, ordered by support (also reported as count of matching patients); confidence is illustrated by a graded scale. Top rules are selected based on the cumulative number of distinct patients associated with at least one rule (90%). Remarkably, all genes of top rules are mildness genes except gene PRSS55, which is present in 55% of female patients.

**Table S1**. Common bi-allelic coding haplotypes of autosomal genes (hetero plus homo versus wt) using the cohort with both genders.

**Table S2**. Common bi-allelic coding haplotypes of autosomal genes (hetero plus homo versus wt) in male cohort.

**Table S3**. Common bi-allelic coding haplotypes of autosomal genes (hetero plus homo versus wt) in female cohort.

**Table S4**. Common bi-allelic coding haplotypes of autosomal genes (homo versus hetero and wt) using the cohort with both genders.

**Table S5**. Common bi-allelic coding haplotypes of autosomal genes (homo versus hetero and wt) in males.

**Table S6**. Common bi-allelic coding haplotypes of X-linked genes (hemy versus wt) in males.

**Table S7**. Rare variants of autosomal genes (hetero plus homo versus wt) using the cohort with both genders.

**Table S8**. Rare variants of autosomal genes (hetero plus homo versus wt) in male cohort.

**Table S9**. Rare variants of autosomal genes (hetero plus homo versus wt) in female cohort. **Table S10**. Rare variants of autosomal genes (homo versus hetero and wt).

**Table S11**. Rare variants of autosomal genes (homo versus hetero and wt) in male cohort.

**Table S12**. Rare variants of autosomal genes (homo versus hetero and wt) in female cohort **Table S13**. Rare variants of X-linked genes (hemy versus wt) in males.

## GEN-COVID Multicenter Study (https://sites.google.com/dbm.unisi.it/gen-covid)

Francesca Montagnani^3,10^, Andrea Tommasi^3,4,5^, Maria Palmieri^3,4^, Gabriele Inchingolo^3,4^, Massimiliano Fabbiani^10^, Barbara Rossetti^10^, Giacomo Zanelli^3,10^, Laura Bergantini^11^, Miriana D’Alessandro^11^, Paolo Cameli^11^, David Bennett^11^, Federico Anedda^12^, Simona Marcantonio^12^, Sabino Scolletta^12^, Federico Franchi^12^, Maria Antonietta Mazzei^13^, Susanna Guerrini^13^, Edoardo Conticini^14^, Luca Cantarini^14^, Bruno Frediani^14^, Danilo Tacconi^15^, Chiara Spertilli^15^, Marco Feri^16^, Alice Donati^16^, Raffaele Scala^17^, Luca Guidelli^17^, Genni Spargi^18^, Marta Corridi^18^, Cesira Nencioni^19^, Leonardo Croci^19^, Gian Piero Caldarelli^20^, Maurizio Spagnesi^21^, Paolo Piacentini^21^, Maria Bandini^21^, Elena Desanctis^21^, Silvia Cappelli^21^, Anna Canaccini^22^, Agnese Verzuri^22^, Valentina Anemoli^22^, Agostino Ognibene^23^, Alessandro Pancrazi^23^, Maria Lorusso^23^, Massimo Vaghi^24^, Antonella D’Arminio Monforte^25^, Esther Merlini^25^, Federica Gaia Miraglia^25^, Mario U. Mondelli^26,27^, Raffaele Bruno^26,27^, Vecchia Marco^26,^, Stefania Mantovani^26^, Serena Ludovisi^26,27^, Massimo Girardis^28^, Sophie Venturelli^28^, Marco Sita^28^, Andrea Cossarizza^29^, Andrea Antinori^30^, Alessandra Vergori^30^, Arianna Emiliozzi^3,10,30^, Stefano Rusconi^31,32^, Matteo Siano^32^, Arianna Gabrieli^32^, Agostino Riva^31,32^, Daniela Francisci^33,34^, Elisabetta Schiaroli^33^, Francesco Paciosi^33^, Pier Giorgio Scotton^35^, Francesca Andretta^35^, Sandro Panese^36^, Stefano Baratti^36^, Renzo Scaggiante^37^, Francesca Gatti^37^, Saverio Giuseppe Parisi^38^, Francesco Castelli^39^, Eugenia Quiros-Roldan^39^, Melania Degli Antoni^39^, Isabella Zanella^40^, Matteo Della Monica^41^, Carmelo Piscopo^41^, Mario Capasso^42,43,44^, Roberta Russo^42,43^, Immacolata Andolfo^42,43^, Achille Iolascon^42,43^, Giuseppe Fiorentino^45^, Massimo Carella^46^, Marco Castori^46^, Giuseppe Merla^46^, Gabriella Maria Squeo^46^, Filippo Aucella^47^, Pamela Raggi^48^, Carmen Marciano^48^, Rita Perna^48^, Matteo Bassetti^49,50^, Antonio Di Biagio^50^, Maurizio Sanguinetti^51,52^, Luca Masucci^51,52^, Serafina Valente^53^, Maria Antonietta Mencarelli^5^, Caterina Lo Rizzo^5^, Chiara Cavalletti^5^, Elena Bargagli^11^, Marco Mandalà^54^, Alessia Giorli^54^, Lorenzo Salerni^54^, Patrizia Zucchi^55^, Pierpaolo Parravicini^55^, Elisabetta Menatti^56^, Tullio Trotta^57^, Ferdinando Giannattasio^57^, Gabriella Coiro^57^, Fabio Lena^58^, Domenico A. Coviello^59^, Cristina Mussini^60^, Giancarlo Bosio^61^, Enrico Martinelli^61^, Sandro Mancarella^62^, Luisa Tavecchia^62^, Lia Crotti^63,64,65,66^, Gianfranco Parati^63,64^, Fausta Sestini^67^, Maurizio Sanarico^68^, Francesco Raimondi^69^, Filippo Biscarini^70^, Alessandra Stella^70^, Marco Rizzi^71^, Franco Maggiolo^71^, Diego Ripamonti^71^, Claudia Suardi^72^, Tiziana Bachetti^73^, Maria Teresa La Rovere^74^, Simona Sarzi-Braga^75^, Maurizio Bussotti^76^, Mattia Bergomi^77^, Katia Capitani^3,78^, Kristina Zguro^3^, Simona Dei^79^, Sabrina Ravaglia^80^

10) Dept of Specialized and Internal Medicine, Tropical and Infectious Diseases Unit, Azienda Ospedaliera Universitaria Senese, Siena, Italy

11) Unit of Respiratory Diseases and Lung Transplantation, Department of Internal and Specialist Medicine, University of Siena, Italy

12) Dept of Emergency and Urgency, Medicine, Surgery and Neurosciences, Unit of Intensive Care Medicine, Siena University Hospital, Italy

13) Department of Medical, Surgical and Neuro Sciences and Radiological Sciences, Unit of Diagnostic Imaging, University of Siena, Italy

14) Rheumatology Unit, Department of Medicine, Surgery and Neurosciences, University of Siena, Policlinico Le Scotte, Italy

15) Department of Specialized and Internal Medicine, Infectious Diseases Unit, San Donato Hospital Arezzo, Italy

16) Dept of Emergency, Anesthesia Unit, San Donato Hospital, Arezzo, Italy

17) Department of Specialized and Internal Medicine, Pneumology Unit and UTIP, San Donato Hospital, Arezzo, Italy

18) Department of Emergency, Anesthesia Unit, Misericordia Hospital, Grosseto, Italy

19) Department of Specialized and Internal Medicine, Infectious Diseases Unit, Misericordia Hospital, Grosseto, Italy

20) Clinical Chemical Analysis Laboratory, Misericordia Hospital, Grosseto, Italy

21) Department of Preventive Medicine, Azienda USL Toscana Sud Est, Italy

22) Territorial Scientific Technician Department, Azienda USL Toscana Sud Est, Italy

23) Clinical Chemical Analysis Laboratory, San Donato Hospital, Arezzo, Italy

24) Chirurgia Vascolare, Ospedale Maggiore di Crema, Italy

25) Department of Health Sciences, Clinic of Infectious Diseases, ASST Santi Paolo e Carlo, University of Milan, Italy

26) Division of Infectious Diseases and Immunology, Fondazione IRCCS Policlinico San Matteo, Pavia, Italy

27) Department of Internal Medicine and Therapeutics, University of Pavia, Italy

28) Department of Anesthesia and Intensive Care, University of Modena and Reggio Emilia, Modena, Italy

29) Department of Medical and Surgical Sciences for Children and Adults, University of Modena and Reggio Emilia, Modena, Italy

30) HIV/AIDS Department, National Institute for Infectious Diseases, IRCCS, Lazzaro Spallanzani, Rome, Italy

31) III Infectious Diseases Unit, ASST-FBF-Sacco, Milan, Italy

32) Department of Biomedical and Clinical Sciences Luigi Sacco, University of Milan, Milan, Italy

33) Infectious Diseases Clinic, Department of Medicine 2, Azienda Ospedaliera di Perugia and University of Perugia, Santa Maria Hospital, Perugia, Italy

34) Infectious Diseases Clinic, “Santa Maria” Hospital, University of Perugia, Perugia, Italy

35) Department of Infectious Diseases, Treviso Hospital, Local Health Unit 2 Marca Trevigiana, Treviso, Italy

36) Clinical Infectious Diseases, Mestre Hospital, Venezia, Italy.

37) Infectious Diseases Clinic, ULSS1, Belluno, Italy

38) Department of Molecular Medicine, University of Padova, Italy

39) Department of Infectious and Tropical Diseases, University of Brescia and ASST Spedali Civili Hospital, Brescia, Italy

40) Department of Molecular and Translational Medicine, University of Brescia, Italy; Clinical Chemistry Laboratory, Cytogenetics and Molecular Genetics Section, Diagnostic Department, ASST Spedali Civili di Brescia, Italy

41) Medical Genetics and Laboratory of Medical Genetics Unit, A.O.R.N. “Antonio Cardarelli”, Naples, Italy

42) Department of Molecular Medicine and Medical Biotechnology, University of Naples Federico II, Naples, Italy

43) CEINGE Biotecnologie Avanzate, Naples, Italy

44) IRCCS SDN, Naples, Italy

45) Unit of Respiratory Physiopathology, AORN dei Colli, Monaldi Hospital, Naples, Italy

46) Division of Medical Genetics, Fondazione IRCCS Casa Sollievo della Sofferenza Hospital, San Giovanni Rotondo, Italy

47) Department of Medical Sciences, Fondazione IRCCS Casa Sollievo della Sofferenza Hospital, San Giovanni Rotondo, Italy

48) Clinical Trial Office, Fondazione IRCCS Casa Sollievo della Sofferenza Hospital, San Giovanni Rotondo, Italy

49) Department of Health Sciences, University of Genova, Genova, Italy

50) Infectious Diseases Clinic, Policlinico San Martino Hospital, IRCCS for Cancer Research Genova, Italy

51) Microbiology, Fondazione Policlinico Universitario Agostino Gemelli IRCCS, Catholic University of Medicine, Rome, Italy

52) Department of Laboratory Sciences and Infectious Diseases, Fondazione Policlinico Universitario A. Gemelli IRCCS, Rome, Italy

53) Department of Cardiovascular Diseases, University of Siena, Siena, Italy

54) Otolaryngology Unit, University of Siena, Italy

55) Department of Internal Medicine, ASST Valtellina e Alto Lario, Sondrio, Italy

56) Study Coordinator Oncologia Medica e Ufficio Flussi Sondrio, Italy

57) First Aid Department, Luigi Curto Hospital, Polla, Salerno, Italy

58) Local Health Unit-Pharmaceutical Department of Grosseto, Toscana Sud Est Local Health Unit, Grosseto, Italy

59) U.O.C. Laboratorio di Genetica Umana, IRCCS Istituto G. Gaslini, Genova, Italy

60) Infectious Diseases Clinics, University of Modena and Reggio Emilia, Modena, Italy

61) Department of Respiratory Diseases, Azienda Ospedaliera di Cremona, Cremona, Italy

62) U.O.C. Medicina, ASST Nord Milano, Ospedale Bassini, Cinisello Balsamo (MI), Italy

63) Istituto Auxologico Italiano, IRCCS, Department of Cardiovascular, Neural and Metabolic Sciences, San Luca Hospital, Milan, Italy

64) Department of Medicine and Surgery, University of Milano-Bicocca, Milan, Italy

65) Istituto Auxologico Italiano, IRCCS, Center for Cardiac Arrhythmias of Genetic Origin, Milan, Italy

66) Istituto Auxologico Italiano, IRCCS, Laboratory of Cardiovascular Genetics, Milan, Italy

67) Department of Medical, Surgical and Neurological Sciences, University of Siena, Italy

68) Independent Data Scientist, Milan, Italy

69) Scuola Normale Superiore, Pisa, Italy

70) CNR-Consiglio Nazionale delle Ricerche, Istituto di Biologia e Biotecnologia Agraria (IBBA), Milano, Italy

71) Unit of Infectious Diseases, ASST Papa Giovanni XXIII Hospital, Bergamo, Italy

72) Fondazione per la ricerca Ospedale di Bergamo, Bergamo, Italy

73) Direzione Scientifica, Istituti Clinici Scientifici Maugeri IRCCS, Pavia, Italy

74) Istituti Clinici Scientifici Maugeri IRCCS, Department of Cardiology, Institute of Montescano, Pavia, Italy

75) Istituti Clinici Scientifici Maugeri, IRCCS, Department of Cardiac Rehabilitation, Institute of Tradate (VA), Italy

76) Cardiac Rehabilitation Unit, Fondazione Salvatore Maugeri, IRCCS, Scientific Institute of Milan, Milan, Italy

77) Veos Digital, Milan, Italy

78) Core Research Laboratory, ISPRO, Florence, Italy

79) Health Management, Azienda USL Toscana Sudest, Tuscany, Italy

80) IRCCS C. Mondino Foundation, Pavia, Italy

## Notes

### Competing Interest Statement

The authors have declared no competing interest.

### Clinical Trial

na

## REFERENCES

1. Islam MR, Hoque MN, Rahman MS, Alam ASMRU, Akther M, Puspo JA, Akter S, Sultana M, Crandall KA, Hossain MA. Genome-wide analysis of SARS-CoV-2 virus strains circulating worldwide implicates heterogeneity. Sci Rep. 2020 Aug 19;10(1):14004.

2. Severe Covid-19 GWAS Group; Ellinghaus, D.; Degenhardt, F.; Bujanda, L.; Buti, M.; Albillos, A.; Invernizzi, P.; Fernández, J.; Prati, D.; Baselli, G. et al. Genomewide Association Study of Severe Covid-19 with Respiratory Failure. N Engl J Med. 2020, 383(16), 1522–1534.

3. Pairo-Castineira, E.; Clohisey, S.; Klaric, L.; Bretherick, A.D.; Rawlik, K.; Pasko, D.; Walker, S.; Parkinson, N.; Fourman, M.H.; Russell, C.D.; et al. Genetic mechanisms of 1. critical illness in Covid-19. Nature. 2020, Dec 11. doi: 10.1038/s41586-020-03065-y. Epub ahead of print. PMID: 3330754.

4. Zhang, X.; Tan, Y.; Ling, Y.; Lu, G.; Liu, F.; Yi, Z.; Jia, X.; Wu, M.; Shi, B.; Xu, S.; et al. Viral and host factors related to the clinical outcome of COVID-19. Nature, 2020;583(7816):437–440.

5. van der Made, C.I.; Simons, A.; Schuurs-Hoeijmakers, J.; van den Heuvel, G.; Mantere, T.; Kersten, S.; van Deuren, R.C.; Steehouwer, M.; van Reijmersdal, S.V.; Jaeger, M.; et al. Presence of Genetic Variants Among Young Men With Severe COVID-19. JAMA. 2020, Jul 24;324(7):1–11.

6. Benetti, E.; Giliberti, A.; Emiliozzi, A.; Valentino, F.; Bergantini, L.; Fallerini, C.; Anedda, F.; Amitrano, S.; Conticini, E.; Tita, R.; et al. Clinical and molecular characterization of COVID-19 hospitalized patients. PLoS One. 2020, Nov 18;15(11):e0242534.

7. Daga, S.; Fallerini, C.; Baldassarri, M.; Fava, F.; Valentino, F;; Doddato, G.; Benetti, E.; Furini, S.; Giliberti, A.; Tita, R.; et al. Employing a systematic approach to biobanking and analyzing clinical and genetic data for advancing COVID-19 research. Eur J Hum Genet. 2021 Jan 17. doi: 10.1038/s41431-020-00793-7. Epub ahead of print. PMID: 33456056.

8. Baldassarri, M.; Picchiotti, N.; Fava, F.; Fallerini, C.; Benetti, E.; Daga S.; Valentino F.; Doddato, G.; Furini, S.; Giliberti, A.; et al. Shorter androgen receptor polyQ alleles protect against life-threatening COVID-19 disease in males. medRxiv 2020.11.04.20225680; doi: https://doi.org/10.1101/2020.11.04.20225680.

9. Fallerini, C.; Daga, S.; Mantovani, S.; Benetti, E.; Pujol, A.; Picchiotti, N.; Schluter, A.; Planas-Serra, L.; Troya, J.; Baldassarri, M.; et al. Association of Toll-like receptor 7 variants with life-threatening COVID-19 disease in males. medRxiv 2020.11.19.20234237; doi: https://doi.org/10.1101/2020.11.19.20234237.

10. Molnar, C. Interpretable-machine-learning. A Guide for Making Black Box Models Explainable”. 2019, https://christophm.github.io/interpretable-ml-book/

11. Picchiotti, N.; Salvioli, M.; Zanardini, E.; Missale, F. COVID-19 pandemic: a mobility-dependent SEIR model with undetected cases in Italy, Europe, and US. Epidemiol Prev, 2020 Sep-Dec;44(5-6 Suppl 2):136–143. English. doi: 10.19191/EP20.5-6.S2.112. PMID: 33412804.

12. Zekavat, S.M.; Lin, S.H.; Bick, A.G.; Liu, A.; Paruchuri, K.; Uddin, M.M.; Ye, Y.; Yu, Z.; Liu, X.; Kamatani, Y.; et al. Hematopoietic mosaic chromosomal alterations and risk for infection among 767,891 individuals without blood cancer. medRxiv [Preprint]. 2020 Nov 16:2020.11.12.20230821. doi: 10.1101/2020.11.12.20230821. PMID: 33236019; PMCID: PMC7685330.

13. Bolton, K.L.; Koh Y.; Foote M.B.;; Im, H.; Jee, J.; Hyun Sun C.; Safonov, A.; Ptashkin, R.; et al. Clonal hematopoiesis is associated with risk of severe Covid-19. http://doi.org/10.1101/2020.11.25.20233163.

