## Supplementary figures and images for "Post-Mendelian genetic model in COVID-19"

### Fig. S1

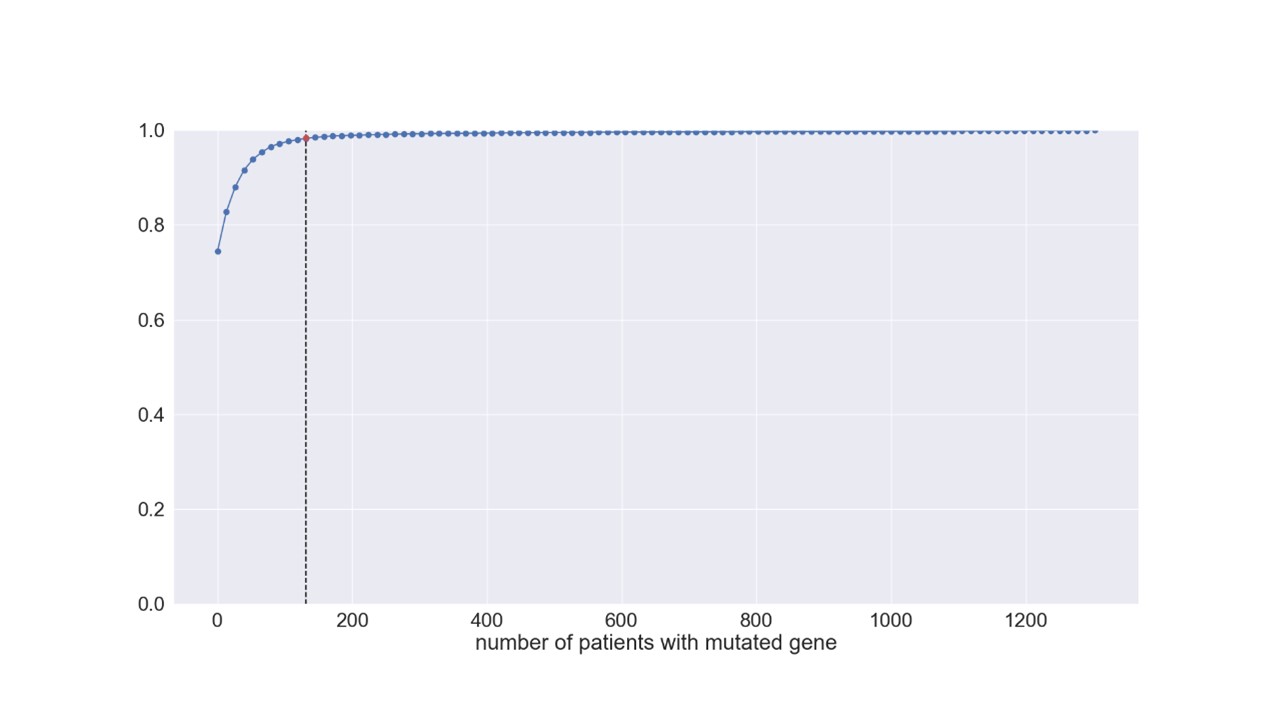

### Figure S2a

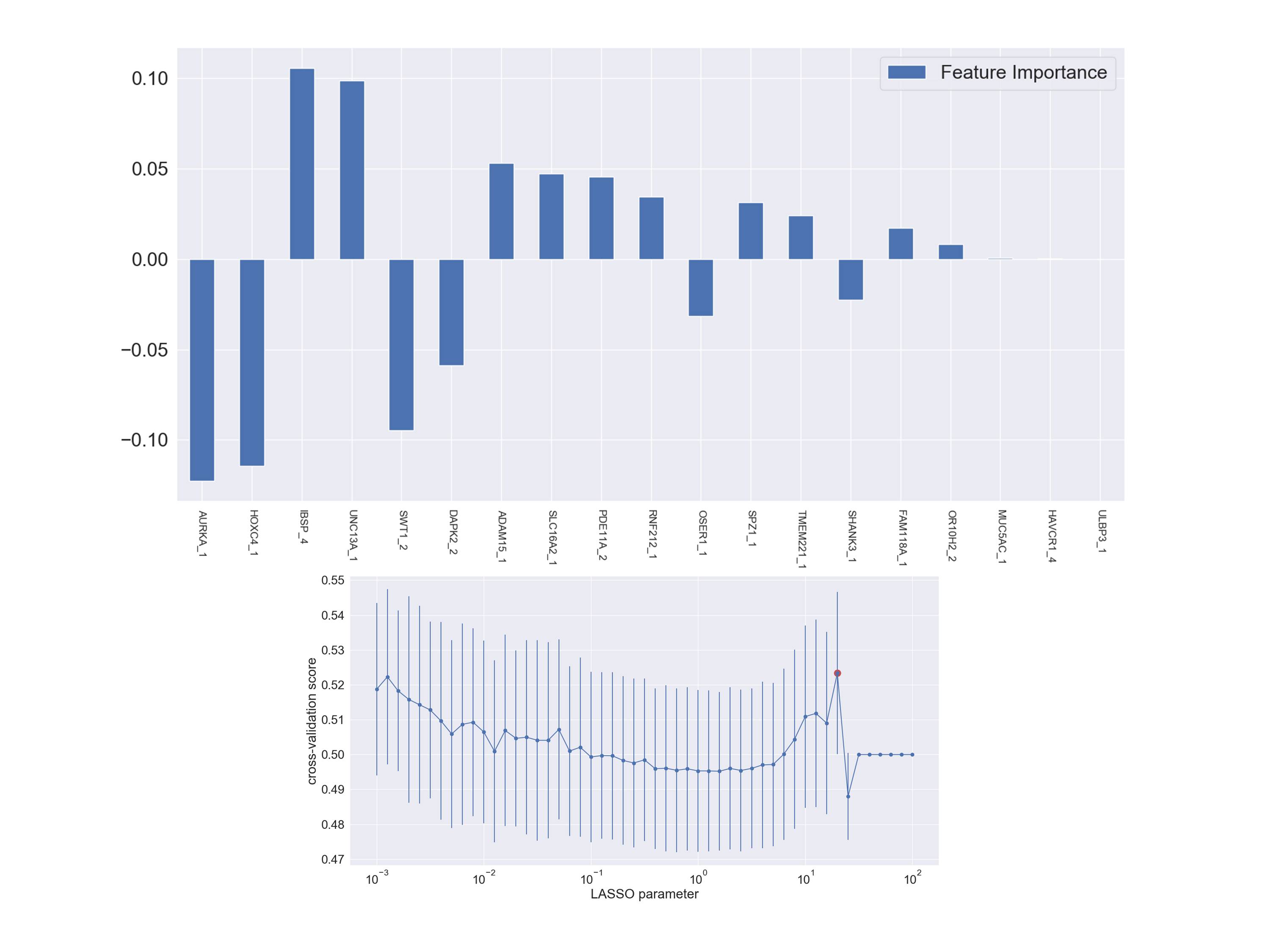

### Figure S2b

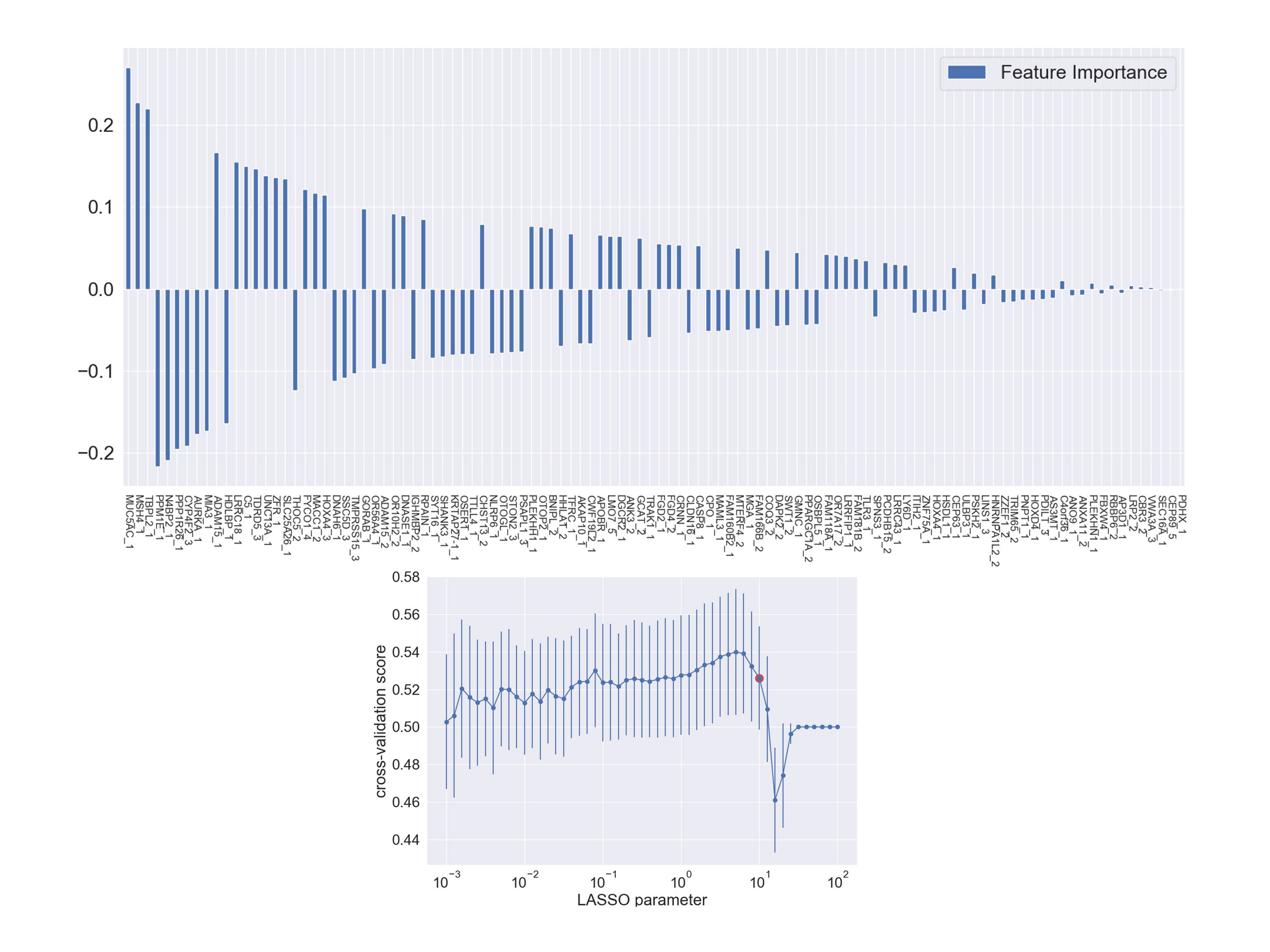

### Figure S2c

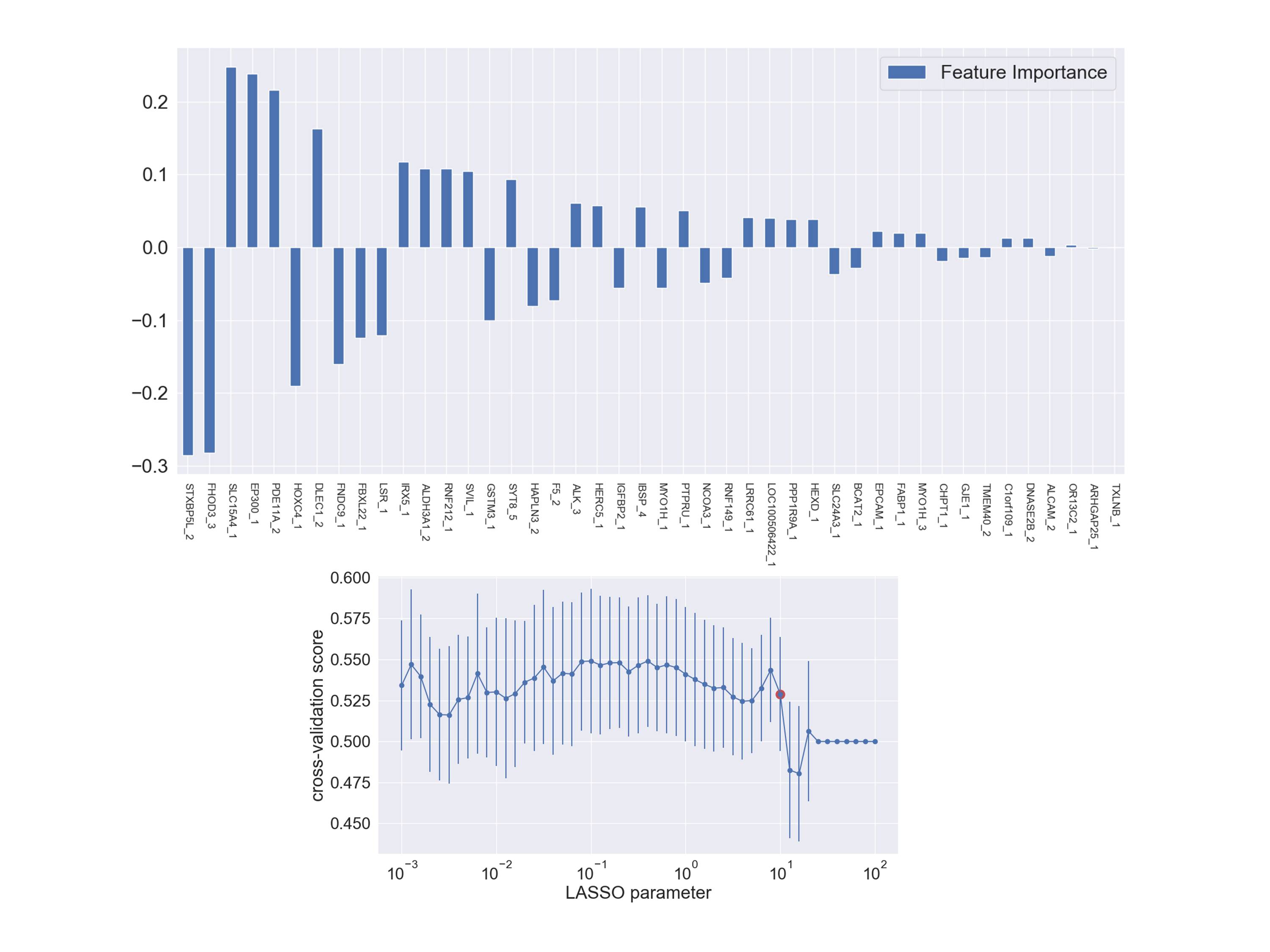

### Figure S2d

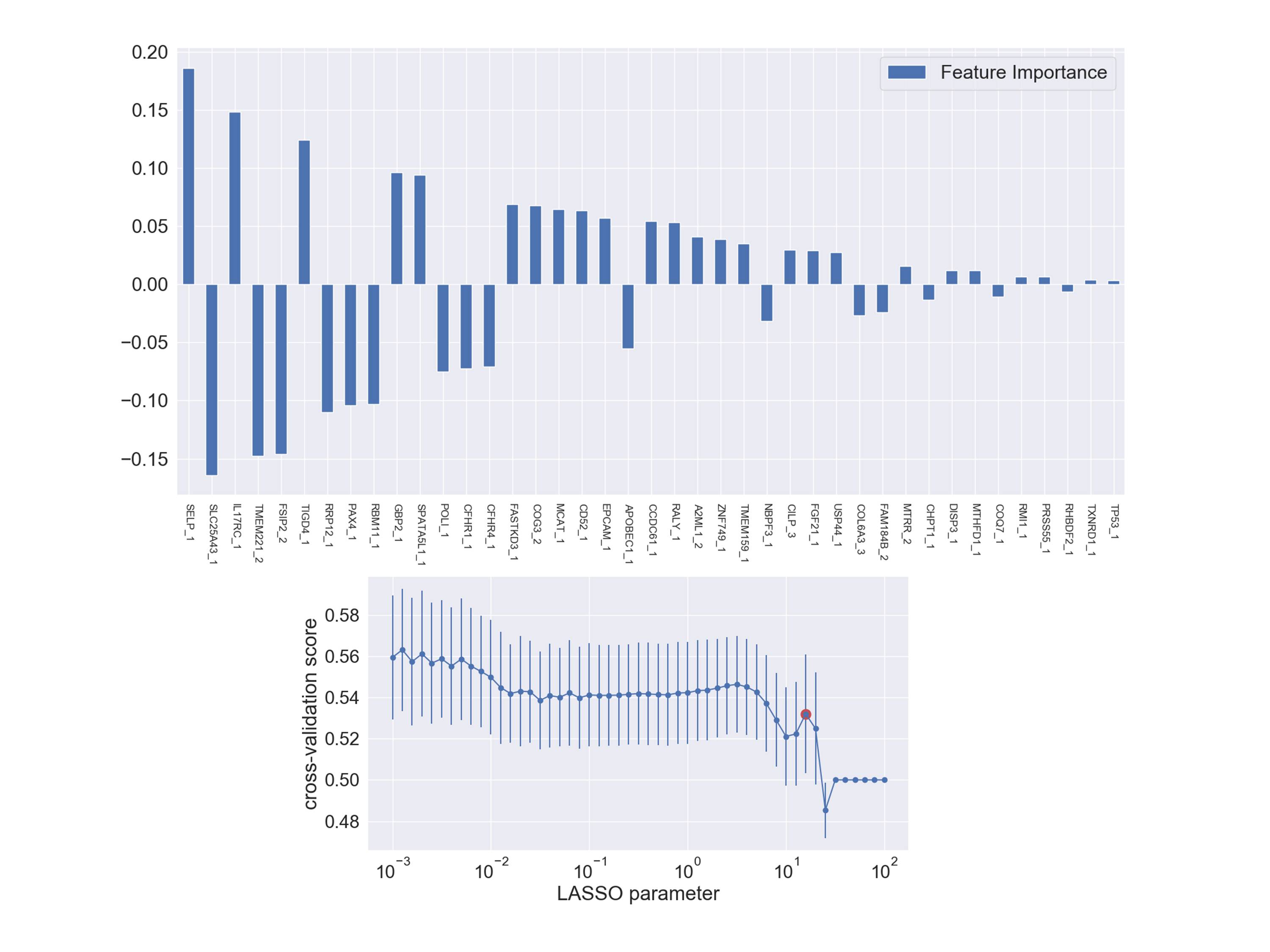

### Figure S2e

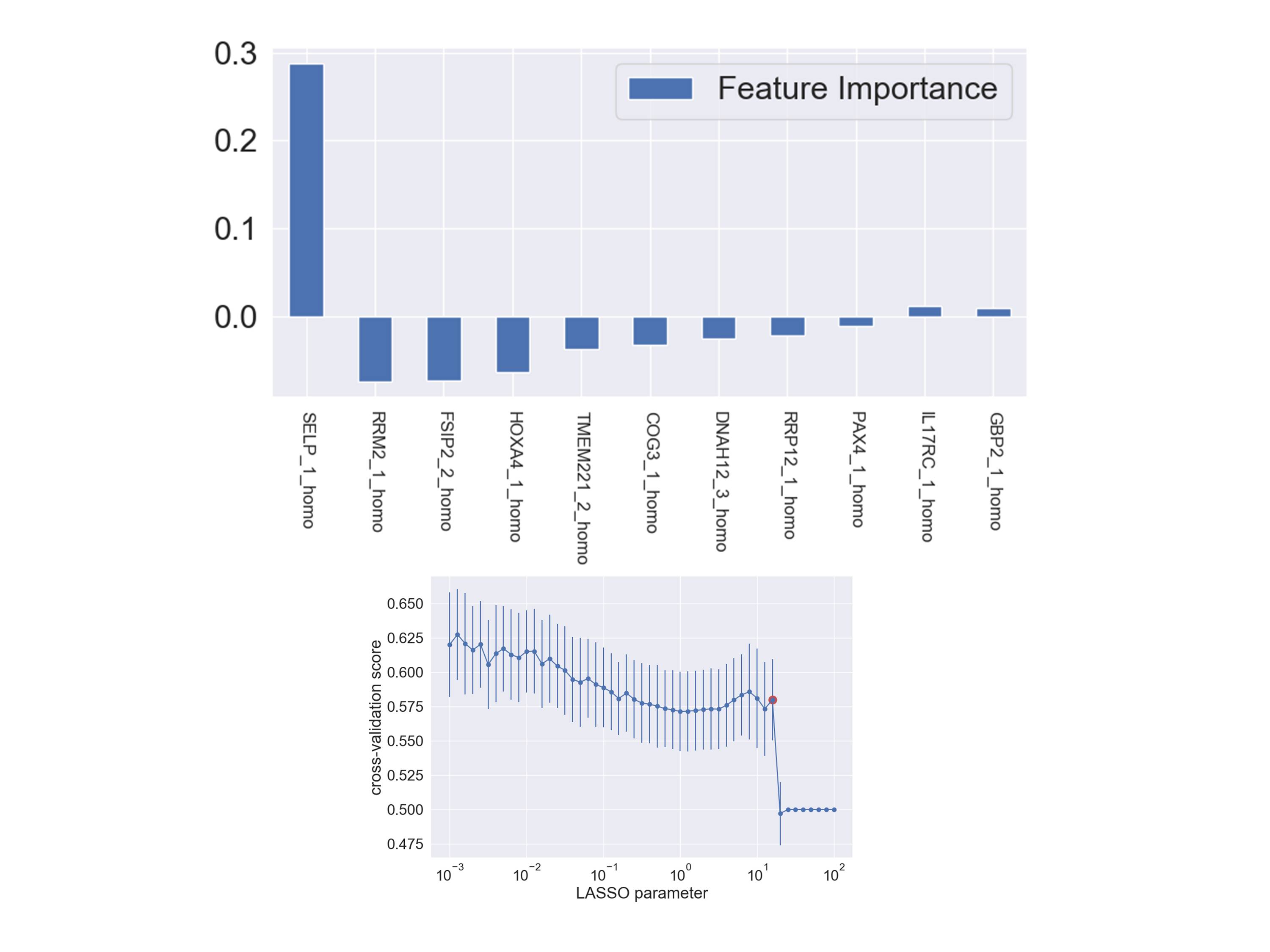

### Figure S2f

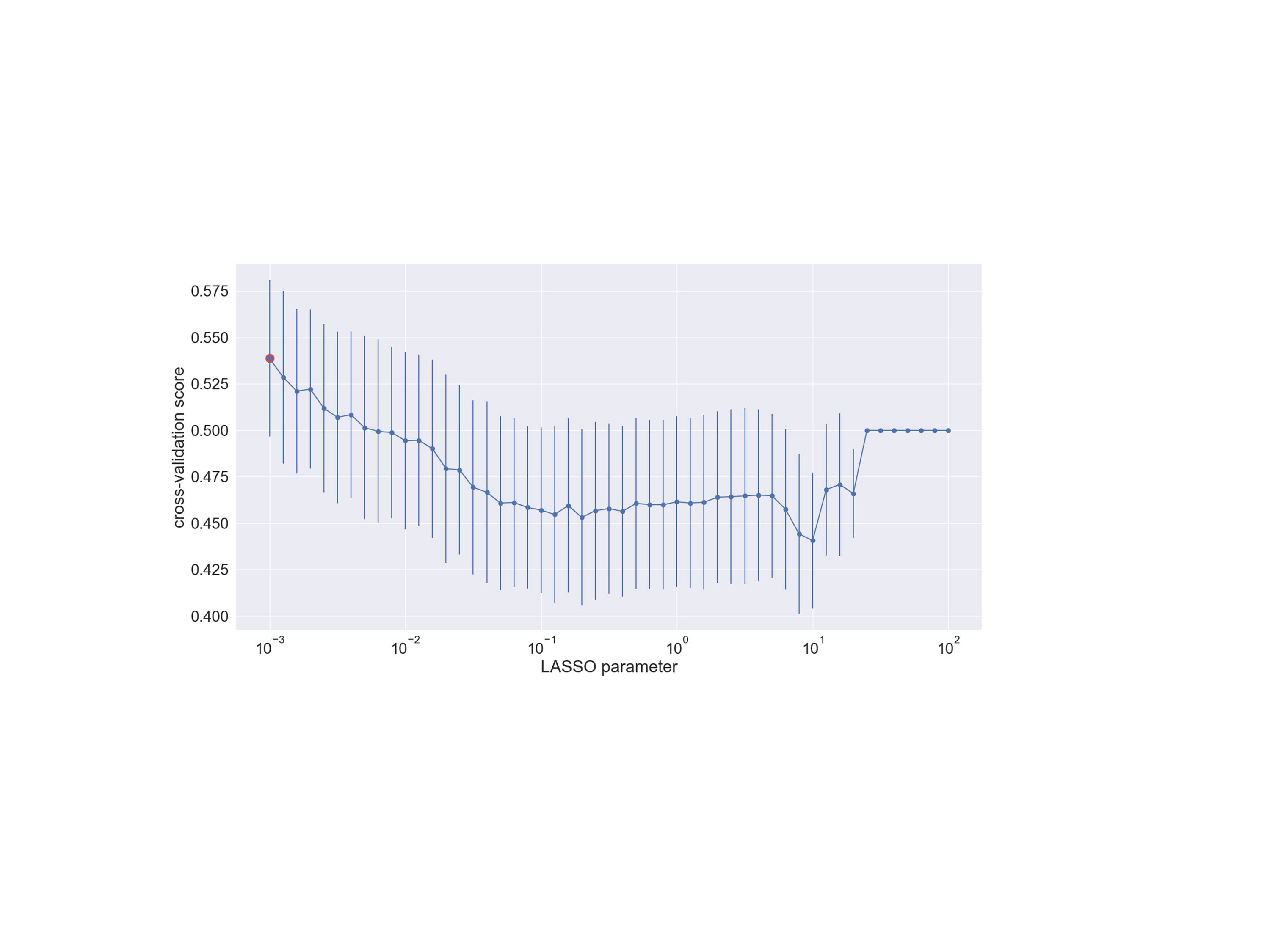

### Figure S2g

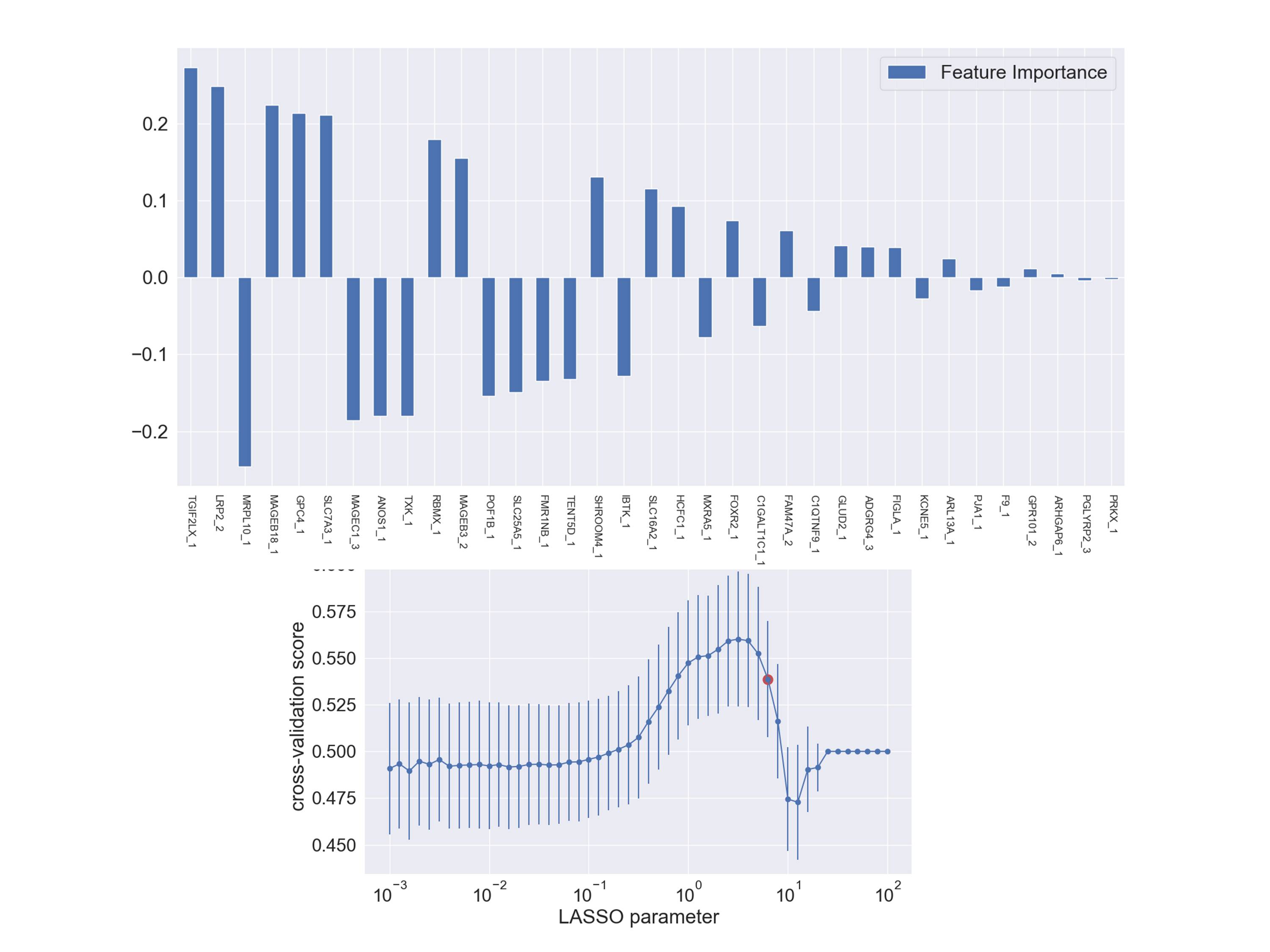

### Figure S2h

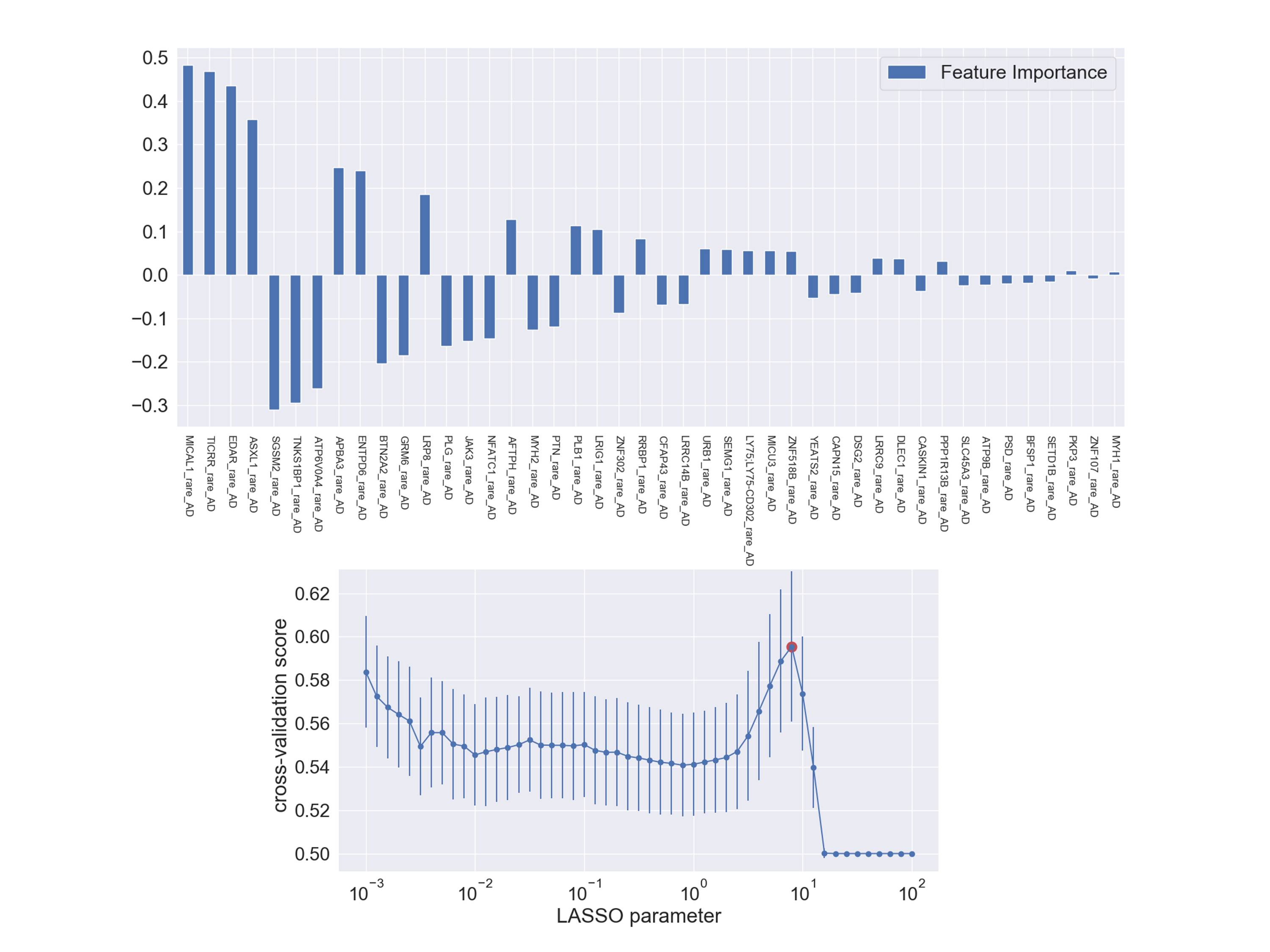

### Figure S2i

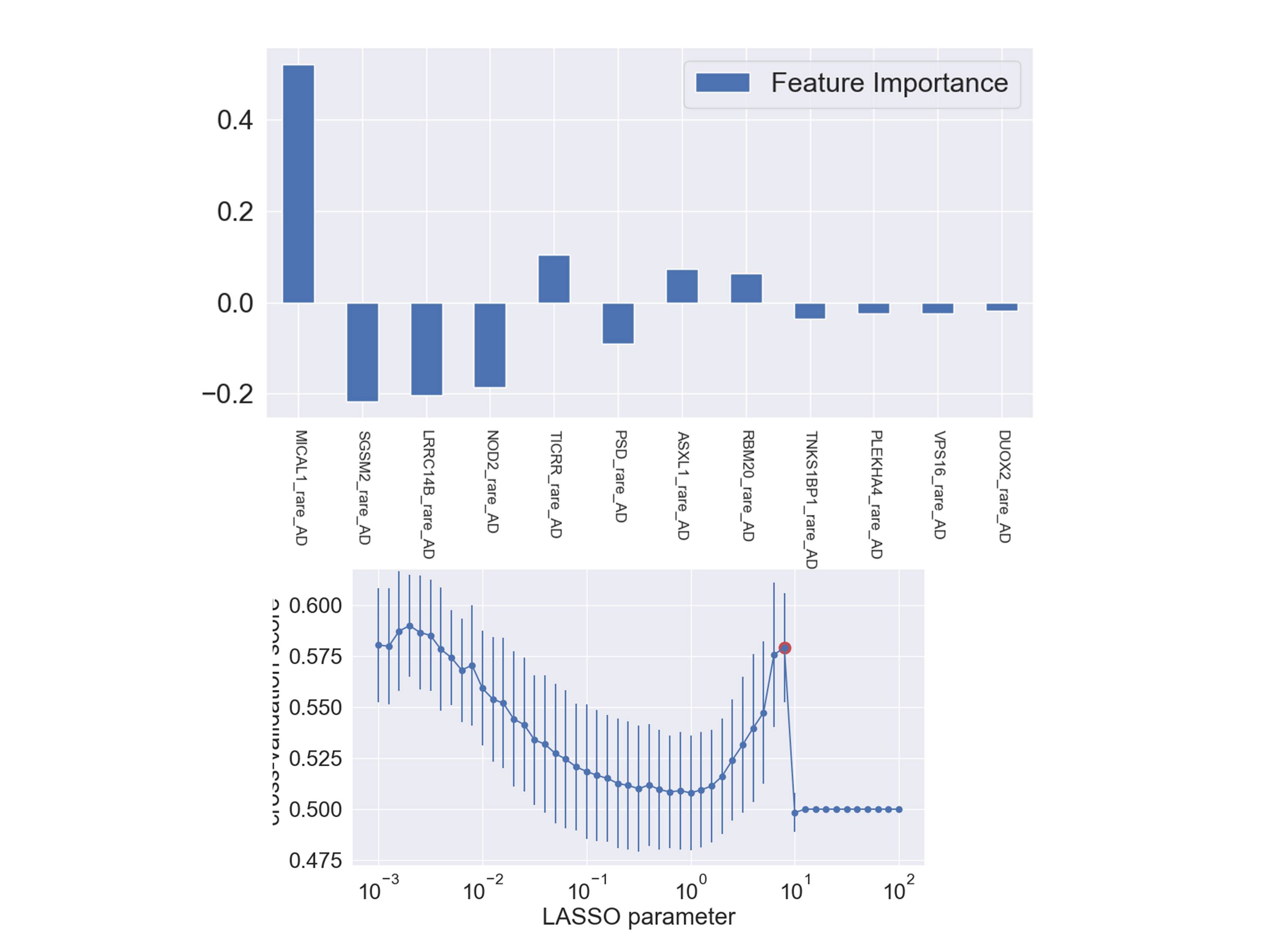

### Figure S2l

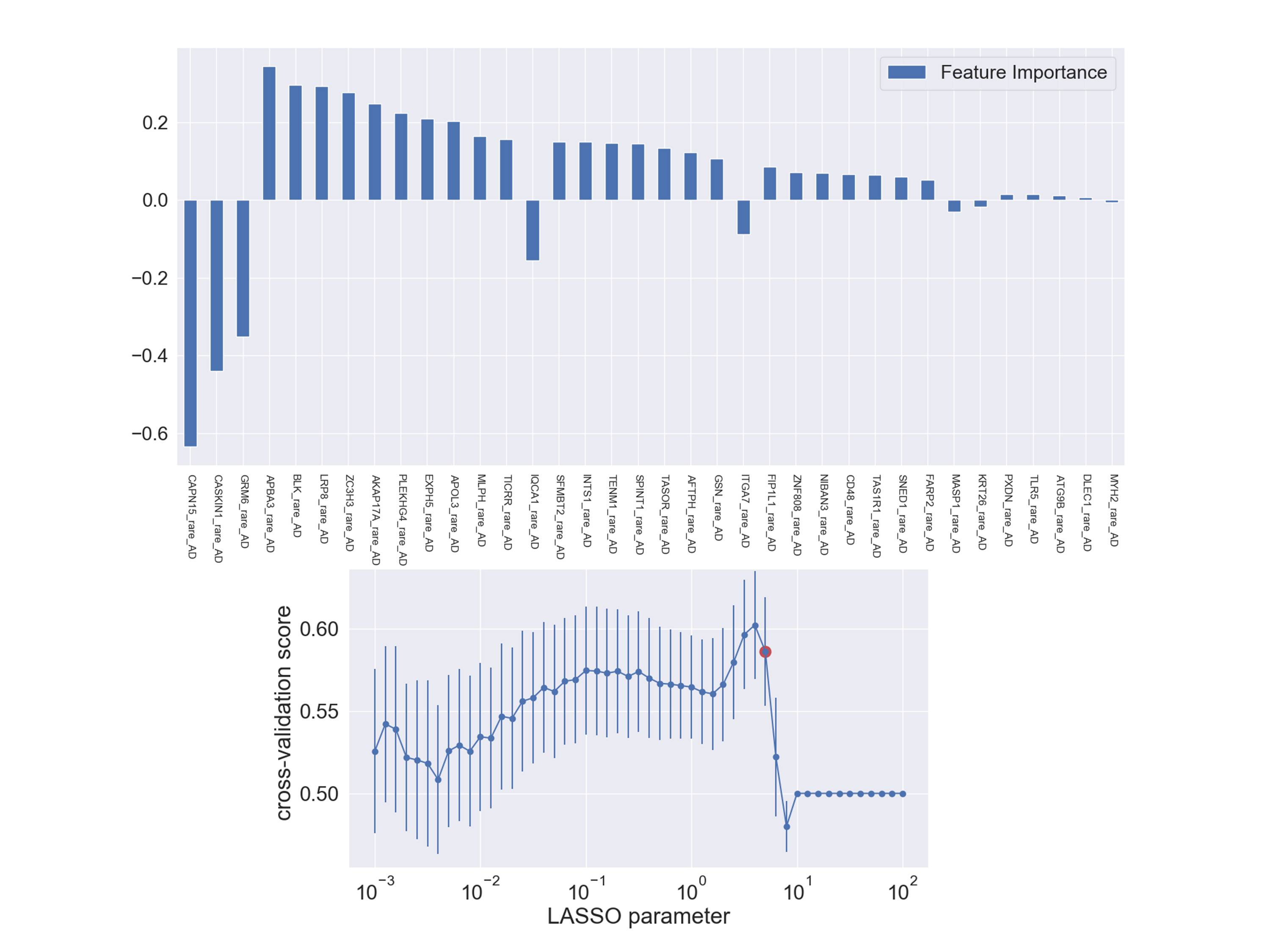

### Figure S2m

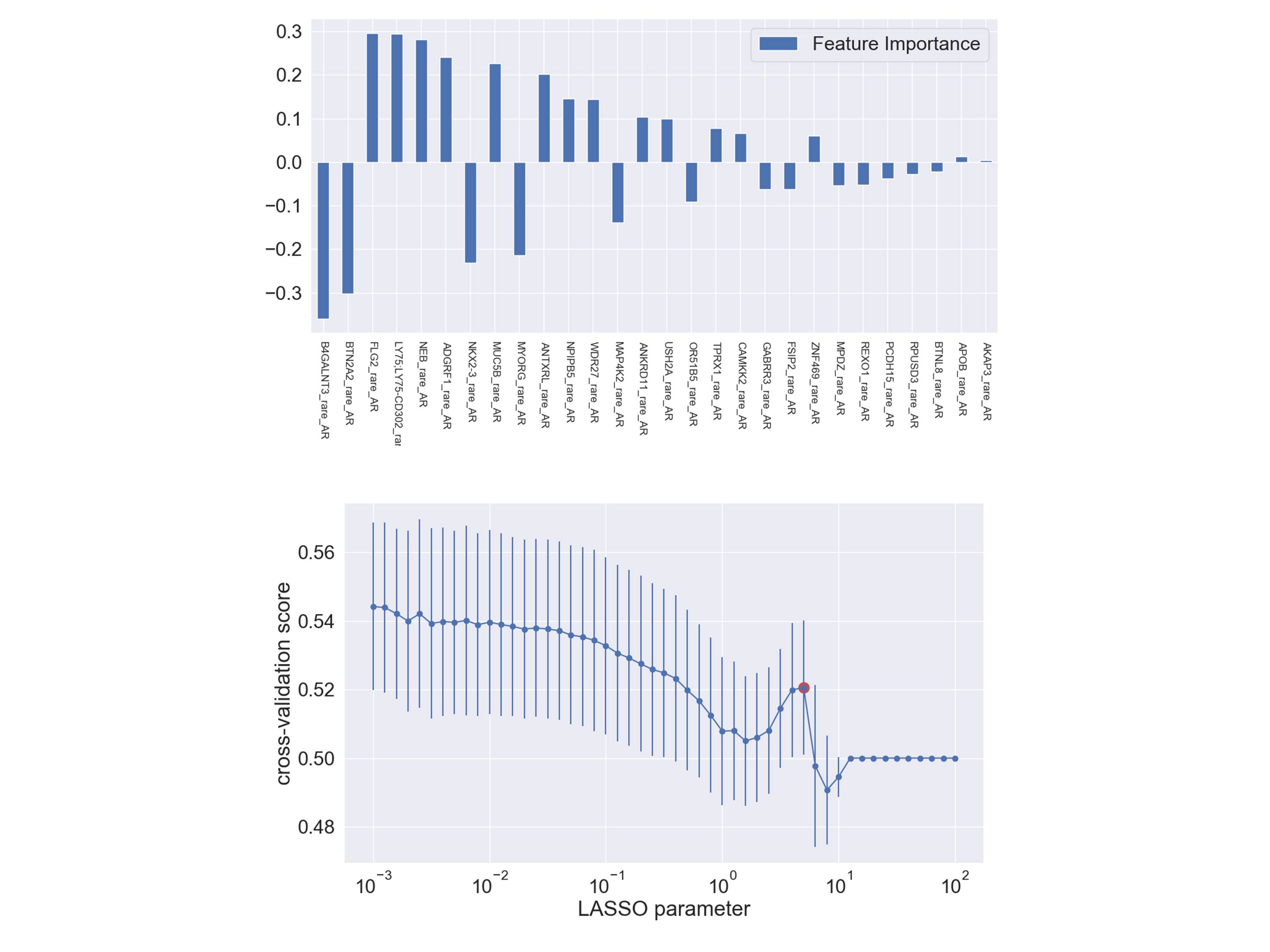

### Figure S2n

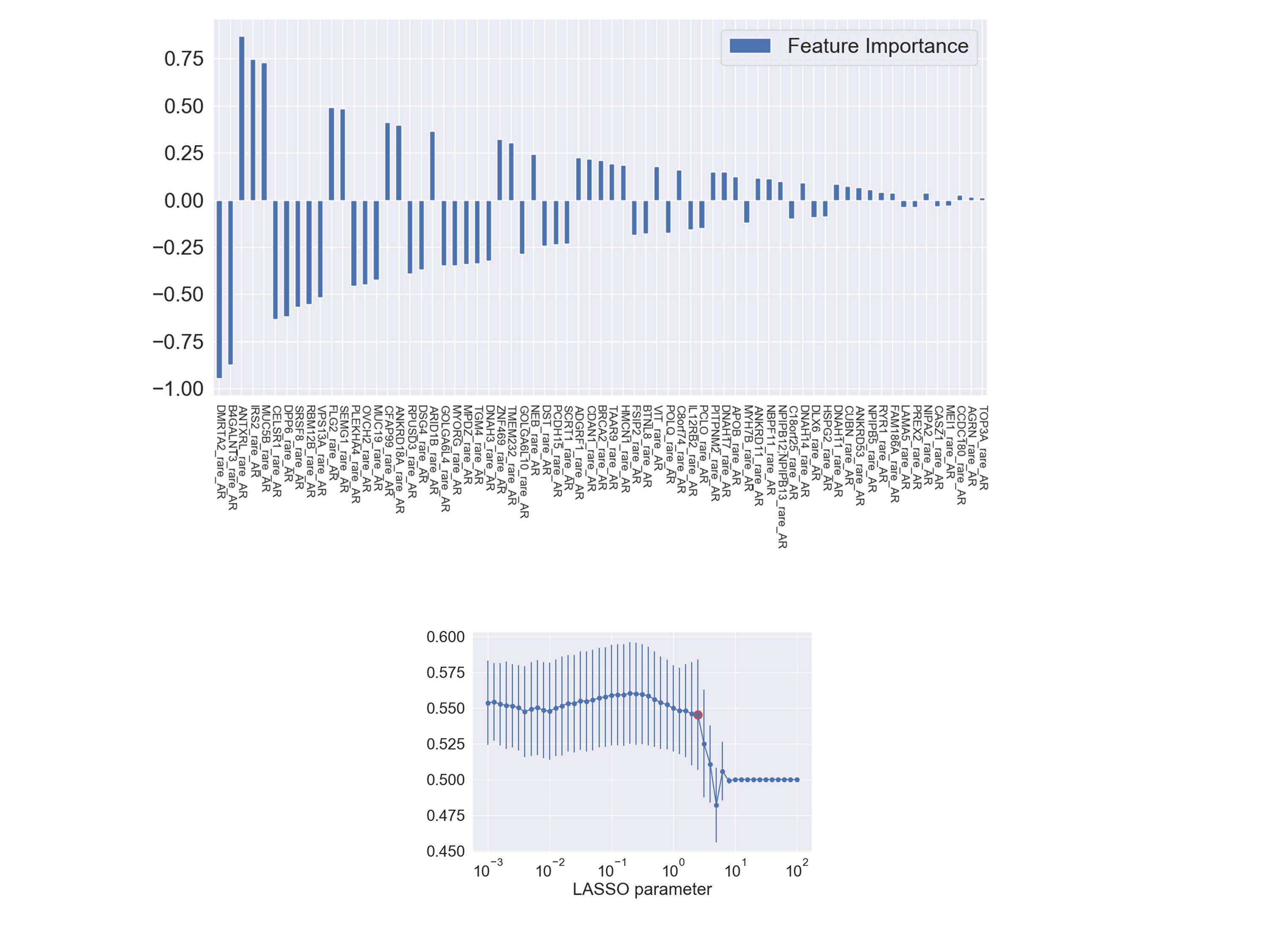

### Figure S2o

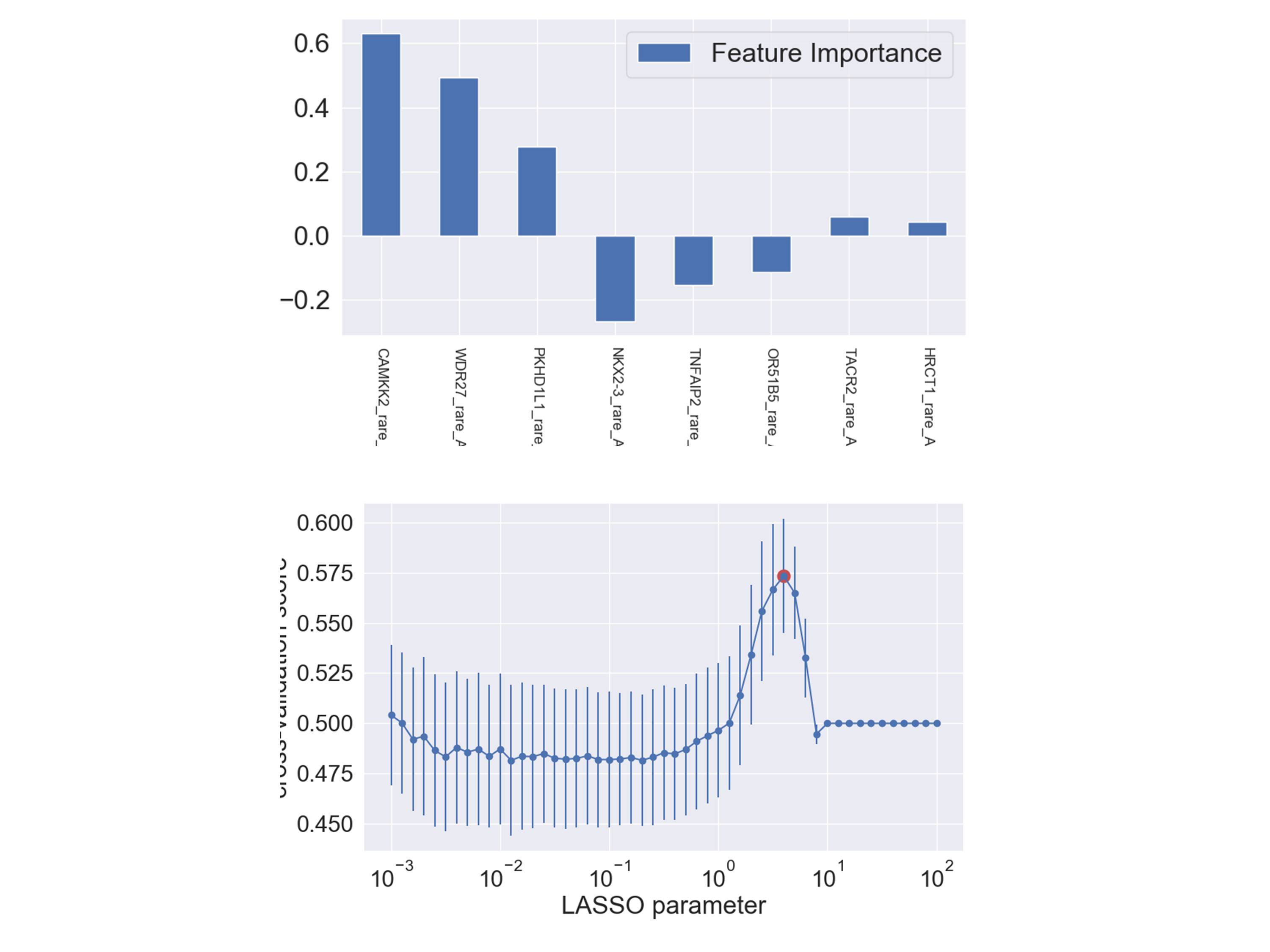

### Figure S2p

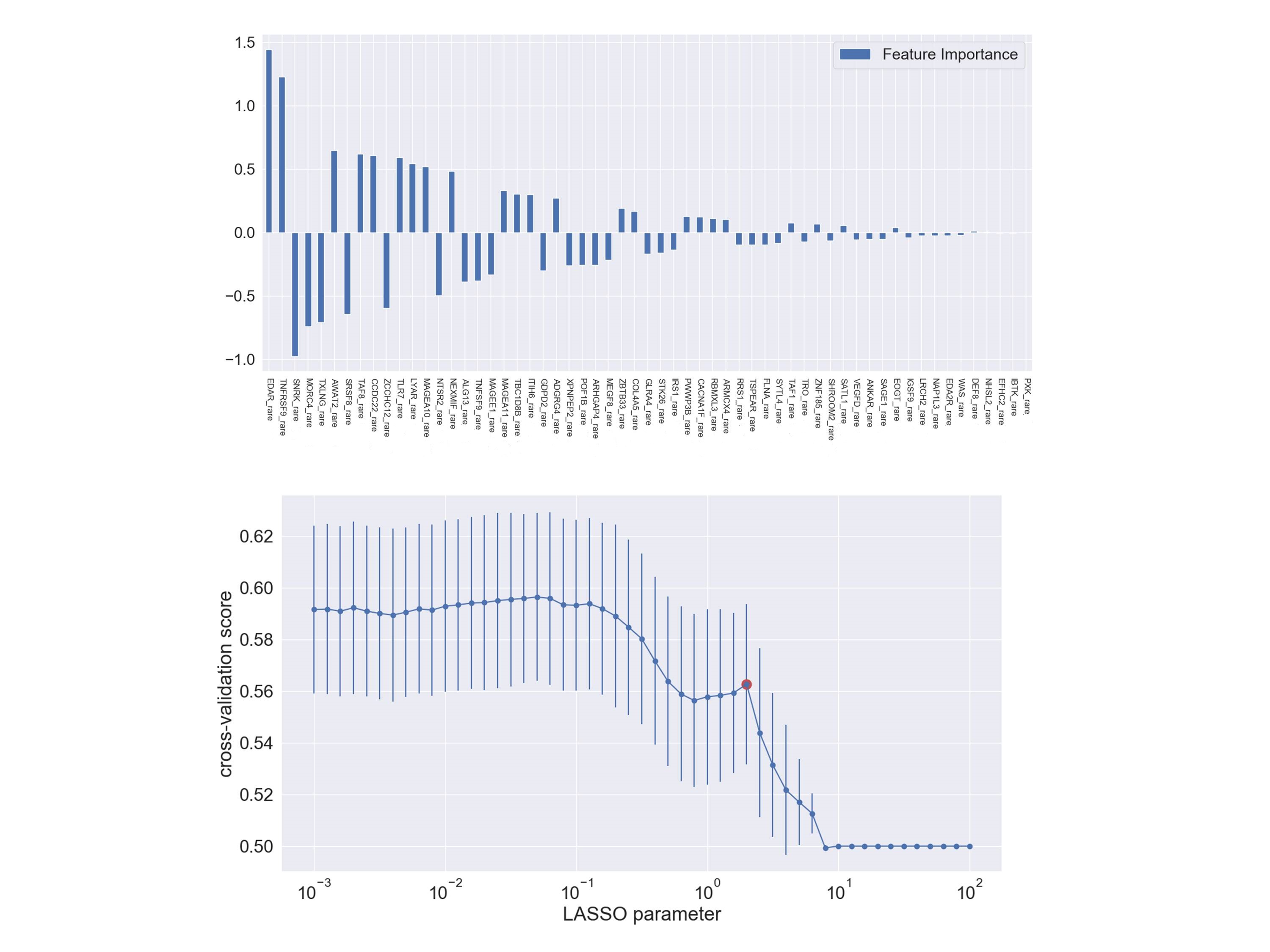

### Figure S3a

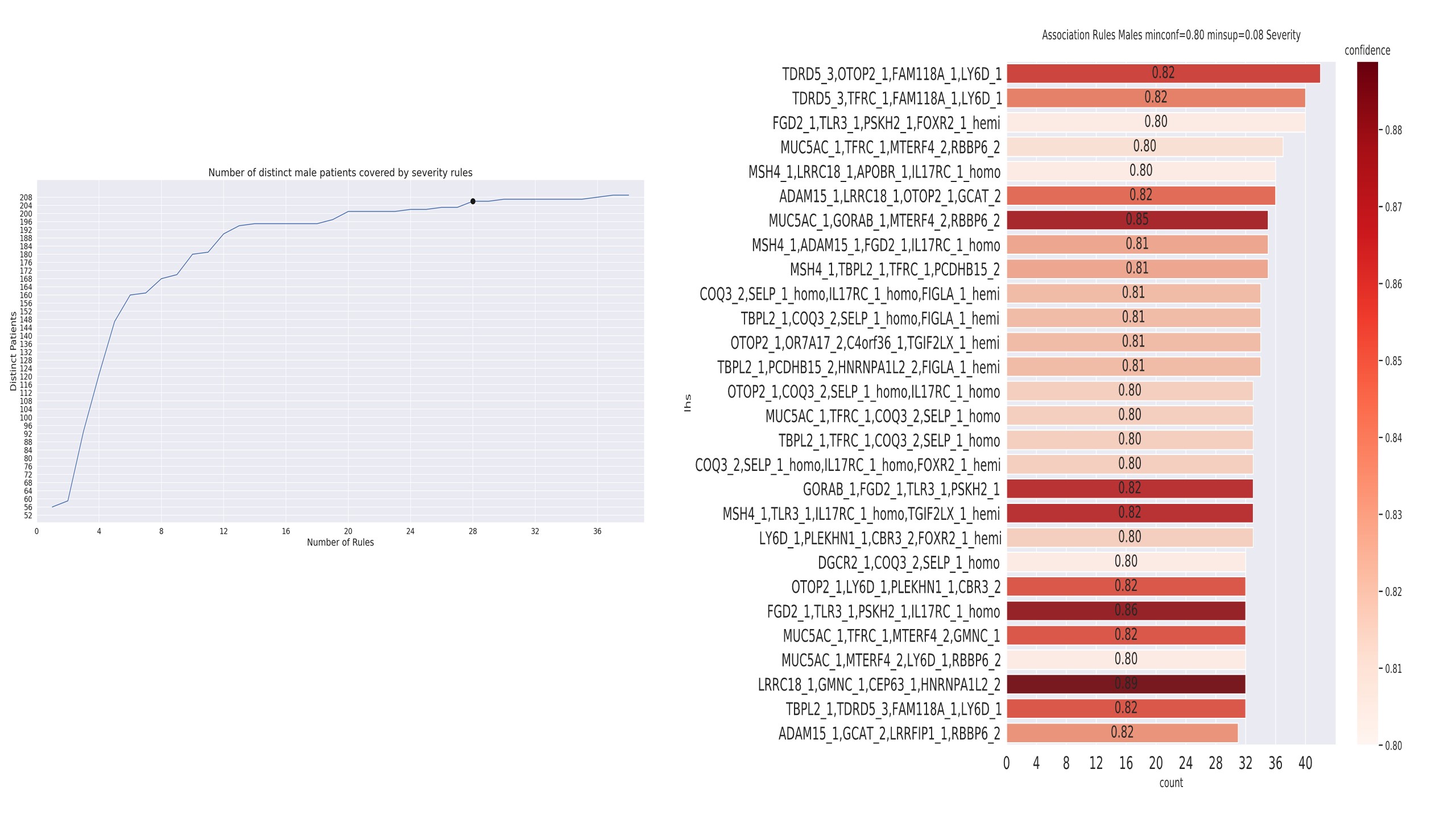

### Figure S3b

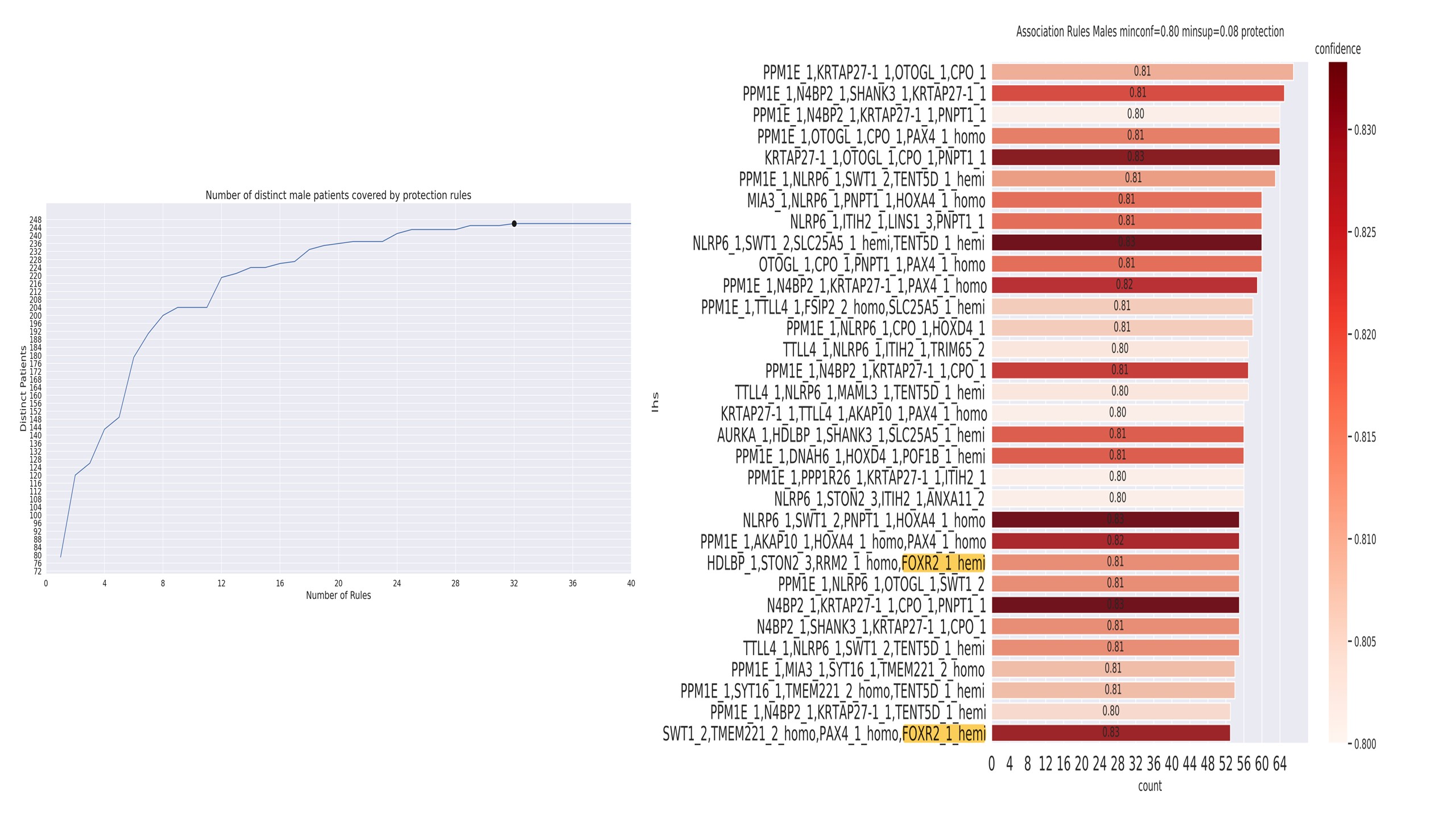

### Figure S3c

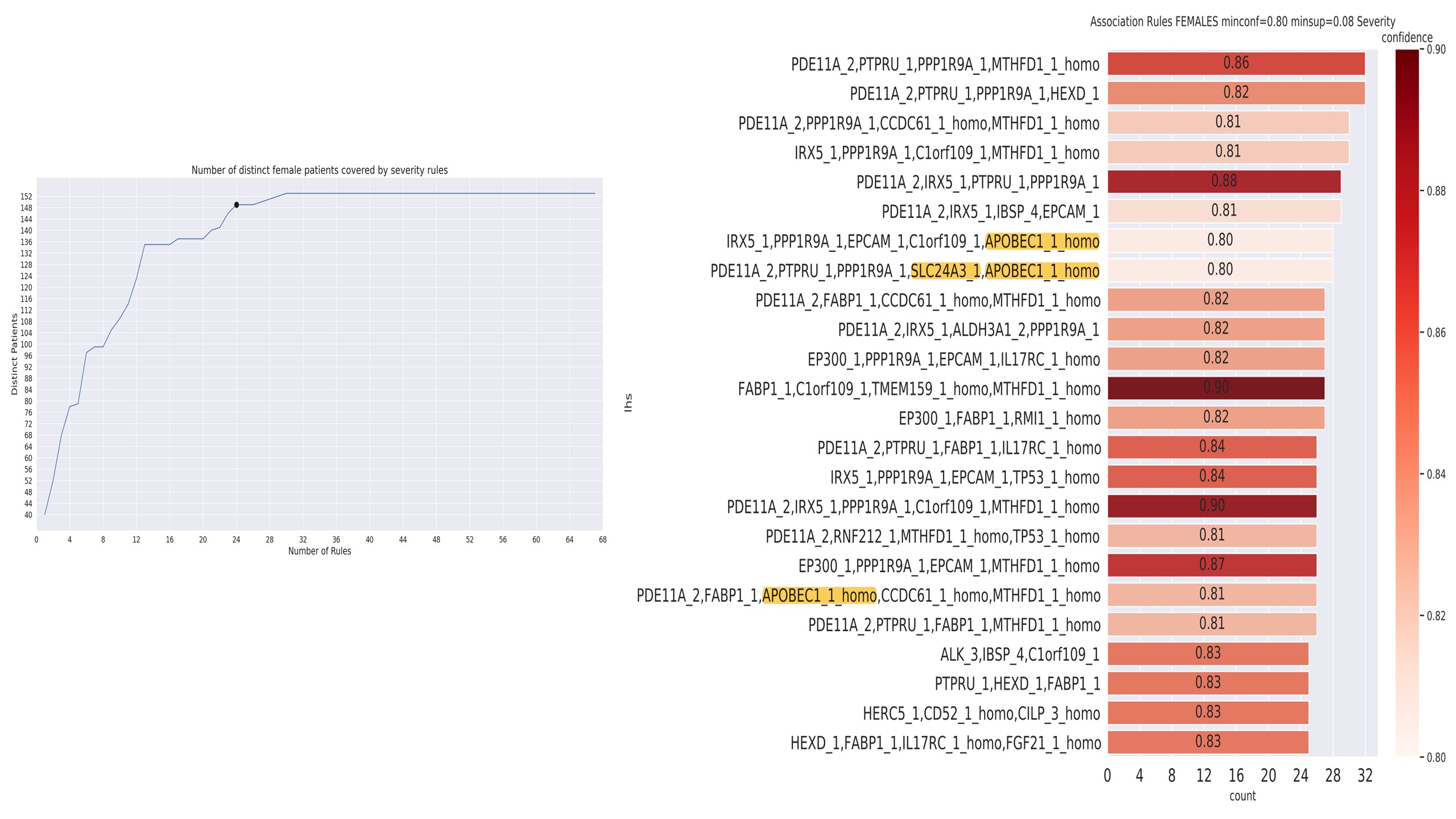

### Figure S3d

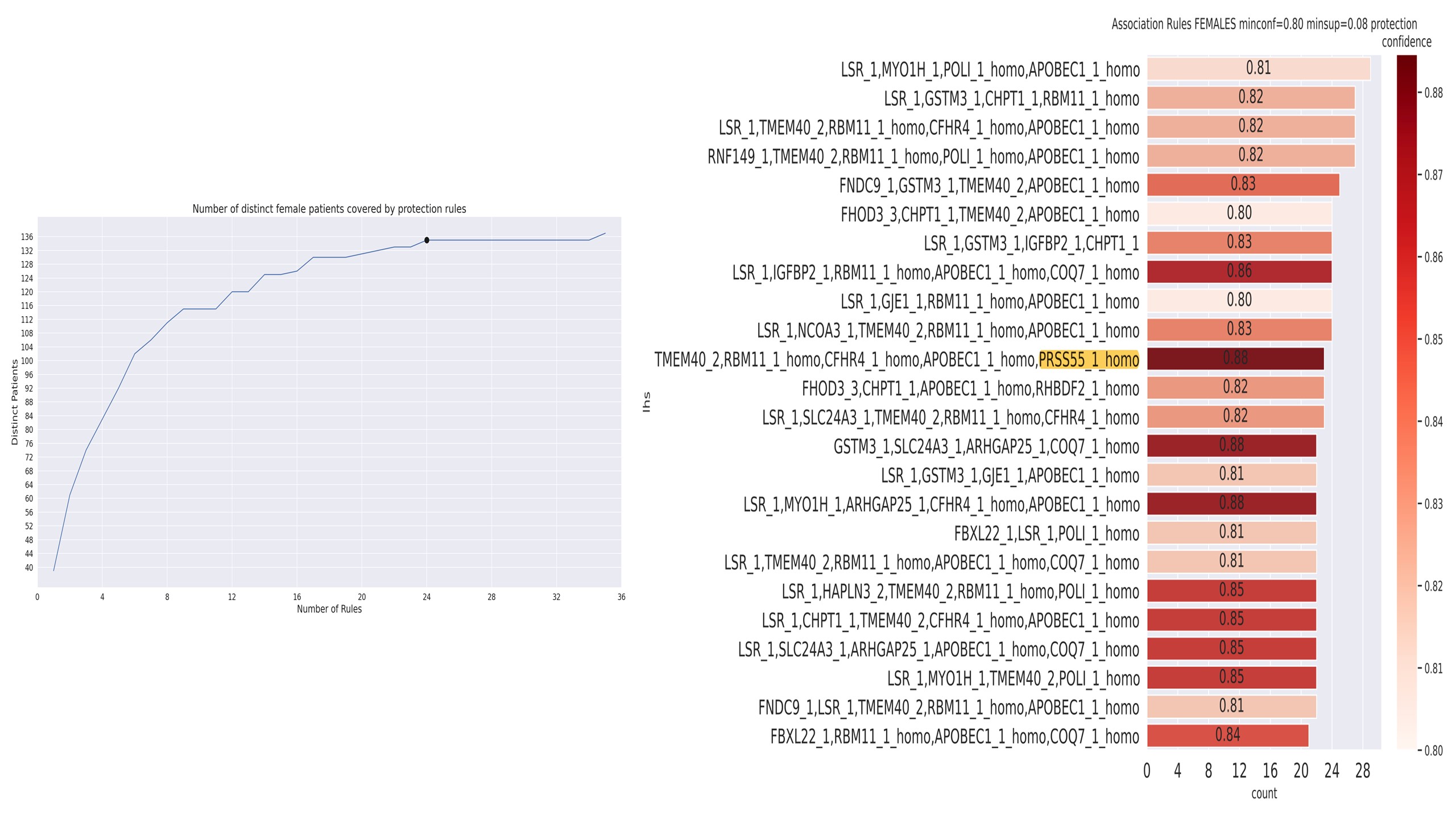
